# Cerebral cortical alterations in adolescent early-onset psychosis: a surface-based morphometry mega-analysis

**DOI:** 10.1101/2025.05.24.25328176

**Authors:** Stener Nerland, Claudia Barth, Kjetil Nordbø Jørgensen, Laura A. Wortinger, Josefina Castro-Fornieles, Covadonga M. Díaz-Caneja, Joost Janssen, Celso Arango, Gisela Sugranyes, Inmaculada Baeza, Fabrizio Piras, Nerisa Banaj, Federica Piras, Simone Elia, Daniela Vecchio, Mathias Lundberg, Hannes Bohman, Bjørn Rishovd Rund, Elena de la Serna, Adriana Fortea, Christian K. Tamnes, Lars T. Westlye, Runar E. Smelror, Dimitrios Andreou, Erik G. Jönsson, Matthew J. Kempton, Shuqing Si, Marinos Kyriakopoulos, Carrie E. Bearden, Kristen Laulette, Saül Pascual-Diaz, Anne-Kathrin J. Fett, Katherine H. Karlsgodt, Lydia Krabbendam, Peter Kochunov, Sophia I. Thomopoulos, Neda Jahanshad, Anthony James, Sophia Frangou, Anne M. Myhre, Nils Eiel Steen, Ole A. Andreassen, Paul M. Thompson, Ingrid Agartz

**Affiliations:** Division for Mental Health and Substance Abuse, Diakonhjemmet Hospital, Oslo, Norway; Division of Mental Health and Addiction, Institute of Clinical Medicine, University of Oslo, Oslo, Norway; Division of Mental Health and Addiction, Vestre Viken Hospital Trust, Drammen, Norway; Department of Psychology, Oslo New University College, Oslo, Norway; Department of Child and Adolescent Psychiatry and Psychology, Hospital Clínic of Barcelona, IDIBAPS, CIBERSAM. Department of Medicine, Insitute of Neuroscience, University of Barcelona, Barcelona, Spain; Department of Child and Adolescent Psychiatry, Institute of Psychiatry and Mental Health, Hospital General Universitario Gregorio Marañón, IiSGM, CIBERSAM, ISCIII, Madrid, Spain; School of Medicine, Universidad Complutense, Madrid, Spain; Department of Child and Adolescent Psychiatry and Psychology, Hospital Clínic of Barcelona, IDIBAPS, CIBERSAM. Barcelona, Spain; Laboratory of Neuropsychiatry, Department of Clinical Neuroscience and Neurorehabilitation, Santa Lucia Foundation IRCCS, Rome, Italy; Department of Clinical Science and Education Södersjukhuset, Karolinska Institutet, Stockholm, Sweden; Department of Neuroscience, Child and Adolescent Psychiatry, Uppsala University, Uppsala, Sweden; Department of Psychology, University of Oslo, Oslo, Norway; Institut d’Investigacions Biomèdiques August Pi I Sunyer (IDIBAPS), Barcelona, Spain; Centro de Investigación Biomédica en Red de Salud Mental (CIBERSAM), Instituto de Salud Carlos III, Madrid, Spain; Adult Psychiatry and Psychology Department, Institute of Neurosciences, Hospital Clínic de Barcelona, Barcelona, Spain; PROMENTA Research Centre, Department of Psychology, University of Oslo, Oslo, Norway; KG Jebsen Centre for Neurodevelopmental Disorders, University of Oslo, Oslo, Norway; Centre for Precision Psychiatry, Division of Mental Health and Addiction, Oslo University Hospital, Oslo, Norway; Centre for Psychiatry Research, Department of Clinical Neuroscience, Karolinska Institutet & Stockholm Health Care Services, Stockholm Region, Stockholm, Sweden; Department of Psychosis Studies, Institute of Psychiatry, Psychology & Neuroscience, King’s College London, London, United Kingdom; 1st Department of Psychiatry, Eginition Hospital, National and Kapodistrian University of Athens, Athens, Greece; Department of Child and Adolescent Psychiatry, Institute of Psychiatry Psychology and Neuroscience, King’s College London, London, UK; South London and Maudsley NHS Foundation Trust, London, UK; Department of Psychiatry and Biobehavioral Sciences, Semel Institute for Neuroscience and Human Behavior, UCLA, Los Angeles, California, USA; Department of Psychology, UCLA, Los Angeles, California, USA; Magnetic Resonance Imaging Core Facility, August Pi i Sunyer Biomedical Research Institute (IDIBAPS), University of Barcelona, Barcelona, Spain; Department of Psychology, School of Health and Medical Sciences, City St George’s, University of London, UK; Department of Psychology, UCLA Los Angeles, California, USA; Department of Psychiatry and Biobehavioral Sciences, UCLA; Department of Clinical, Neuro, and Developmental Psychology, Institute for Brain and Behavior Amsterdam, Vrije Universiteit Amsterdam, the Netherlands; Department of Psychiatry and Behavioral Sciences, The University of Texas Health Science Center Houston, Houston, TX, USA; Imaging Genetics Center, Mark & Mary Stevens Neuroimaging & Informatics Institute, Keck School of Medicine, University of Southern California, Marina del Rey, CA, USA; Department of Psychiatry, University of Oxford, Oxford, UK; Highfield Unit, Warneford Hospital, Oxford, UK; Department of Psychiatry, Icahn School of Medicine at Mount Sinai, New York, New York, USA; Section of Child and Adolescent Mental Health Research, Division of Mental Health and Addiction, Oslo University Hospital, Oslo, Norway; Institute of Clinical Medicine, University of Oslo, Oslo, Norway

## Abstract

Cortical brain morphology in early-onset psychosis (EOP; age of onset < 19 years) is poorly understood, partly due to recruitment constraints linked to its low incidence. We pooled T1-weighted magnetic resonance imaging (MRI) data from 387 adolescents with EOP (mean age=16.1±1.5; 49.6% female) and 338 healthy controls (CTR; mean age=15.8±1.9, 54.4% female) from nine research sites worldwide. Using harmonized processing protocols with FreeSurfer, we extracted cortical brain metrics from 34 bilateral regions. Univariate regression analysis revealed widespread lower bilateral cortical thickness (left/right hemisphere: *d*=-0.36/-0.31), surface area (left/right: *d*=-0.42/-0.41), cortical volume (left/right: *d*=-0.58/-0.56), and Local Gyrification Index (LGI; left/right: *d*=-0.39/-0.52) in EOP relative to CTR. Subgroup analyses showed broader and more pronounced case-control differences in early-onset schizophrenia for area, volume, and LGI. We found no associations with antipsychotic medication use, illness duration, age of onset, or positive symptoms. Negative symptoms were related to smaller left lingual volumes (partial *r*=-0.21; *p_FDR_*=0.014) and antidepressant users had smaller area (*d*=-0.43; *p_FDR_*=0.034) and volume (*d*=-0.50; *p_FDR_*=0.003) of the right rostral anterior cingulate compared to non-users. Cortical alterations in EOP showed a similar pattern to those observed in prior studies on adults with schizophrenia (SCZ; *r*=0.62) and bipolar disorders (BD; *r*=0.61). However, surface area alterations were overall 1.5 times greater for EOP than adult SCZ and 4.6 times greater than adult BD. In the largest study of its kind, we observed an extensive pattern of cortical alterations in adolescents with psychotic disorders, highlighting the potential impact of aberrant neurodevelopment on cortical morphology in this clinical group.

## 1. Introduction

The morphology of the cerebral cortex has been extensively studied in adults with psychotic disorders [1,2]. Less is known about morphological brain alterations in adolescents with early-onset psychosis (EOP), i.e., psychotic disorders diagnosed before 19 years of age [3]. This knowledge gap is compounded by the relatively low incidence of EOP (∼0.6%), making the recruitment of large samples challenging and thus limiting the statistical sensitivity and robustness required for state-of-the-art neuroimaging analyses. Moreover, EOP encompasses a heterogeneous clinical spectrum, including early-onset schizophrenia (EOS), affective psychosis (AFP) and other non-affective psychotic disorders (OTP) [4]. This clinical heterogeneity necessitates large sample sizes to enable subgroup analyses. We therefore formed the EOP Working Group within the Enhancing NeuroImaging Genetics Through Meta Analysis consortium (ENIGMA; http://enigma.ini.usc.edu) [5] to facilitate international collaboration between research groups collecting magnetic resonance imaging (MRI) and clinical data on adolescents with EOP.

Psychotic disorders most commonly emerge in early adulthood [6,7], with an estimated lifetime prevalence of approximately 3% [8]. However, the incidence during adolescence is lower, with one Danish nationwide population-based cohort study estimating a cumulative incidence of 0.76% in girls and 0.48% in boys before the age of 18 [9]. Despite its rarity, EOP is a leading contributor to global disease burden [10], driven by its prolonged course, severe and lasting functional impairment, and disruption of critical social and educational milestones during a critical developmental period [11]. Earlier psychosis onset is associated with worse long-term outcomes [12], with EOS linked to a poorer prognosis than adult-onset schizophrenia (SCZ) [13]. An adolescent onset of a psychotic disorder coincides with critical brain maturational processes, hypothesized to heighten vulnerability to structural and functional brain alterations [14] and reflect greater neurodevelopmental involvement than in adults with SCZ [14,15]. Specifically, EOS may exhibit more pronounced neuroanatomical alterations, reflecting greater illness severity and developmental and functional impairments compared to AFP and OTP [16,17]. There is an urgent need to advance our understanding of cortical brain alterations in EOP, their associations with clinical characteristics, and how they compare to those observed in adult patients with SCZ.

Prior MRI studies of the cerebral cortex have reported lower cortical thickness and cerebral grey matter (GM) volume in children and adolescents with psychotic disorders compared to healthy controls [14,15,18–22]. In a previous ENIGMA-EOP voxel-based morphometry (VBM) study, we detected a pattern of lower regional GM volumes across most of the cerebral cortex [23], which were most pronounced in the cingulate and the frontal and temporal cortices. Furthermore, lower volumes in the cerebellum, thalamus, and left inferior parietal gyrus were associated with older age of onset. These findings are consistent with prior studies of adolescents with EOP, although alterations were greater than those reported in another meta-analysis of brain abnormalities in EOS [24]. Early longitudinal studies of childhood-onset schizophrenia (COS; onset < 12 years) have reported a fourfold greater decline in cortical GM volume [25], as well as accelerated GM volume loss progressing from parietal to temporal and prefrontal regions [26], in COS compared to healthy controls. More recently, Pina-Camacho et al. (2022) reported accelerated longitudinal cortical thinning in adolescent-onset, but not adult-onset, first-episode psychosis patients [27], indicating more pronounced longitudinal cortical thinning in adolescents with EOP [28].

Most previous studies of cerebral cortical morphology in EOP have employed a VBM approach, whereas few studies to date have used surface-based morphometry (SBM) [18,29,30]. Previous studies suggest that SBM offers enhanced sensitivity to pathological alterations in certain clinical conditions [31] and tissue types, notably the cerebral cortex [32]. Importantly, SBM distinguishes between cortical thickness and surface area, two imaging phenotypes that together contribute to cortical GM volume, but are thought to reflect distinct genetic, neurobiological, and maturational processes [33–36]. In the present study, we used an SBM approach based on FreeSurfer [37,38], which also enables direct comparisons with prior large-scale ENIGMA studies of cortical thickness and surface area in adults with SCZ [39] and bipolar disorders (BD) [40]. Moreover, surface-based approaches allow us to quantify the degree of brain gyrification, e.g., using the Local Gyrification Index (LGI) [41]. Such cortical folding indices are of particular interest in the study of disorders with a known or suspected neurodevelopmental aetiology, since cortical folding is largely established prenatally and gyrification indices are therefore thought to reflect early neurodevelopmental processes [42]. Indeed, prior studies of adolescent psychosis have reported differences in cortical folding in EOP relative to healthy controls [43,44]; however, these studies have been constrained by small sample sizes that limit statistical power.

Here, we conducted the first large-scale mega-analysis of cortical thickness, surface area, cortical volume, and LGI in 387 adolescents with EOP and 338 age-matched healthy controls from nine research sites worldwide. We hypothesized thinner thickness, particularly in the cingulate, temporal, and frontal cortices, smaller global surface area, and lower LGI, particularly in the left precentral gyrus, right middle temporal gyrus, and the right precuneus [45]. In diagnostic subgroup analyses, we hypothesized greater cortical alterations in EOS than AFP or OTP. To probe the hemispheric specificity of case-control differences, we investigated the cortical asymmetry index [46,47]. Furthermore, we compared the pattern of cortical morphology alterations in adolescents with those of adults with SCZ and BD. We hypothesized that surface area and LGI would be more severely affected in adolescents with EOP. For all cortical metrics, we assessed sex differences and the impact of antipsychotic and antidepressant medication use, psychotic symptom severity, age of onset, and duration of illness. Given the paucity of large-scale studies of cortical morphology in EOP, these analyses were exploratory in nature. Finally, to assess cross-site heterogeneity, we conducted a supplementary meta-analysis.

## 2. Materials & methods

### Study sample

The mega-analysis dataset comprised 387 individuals with EOP (49.4% female) and 338 healthy controls (CTR; 54.4% female) recruited at nine research sites participating through the ENIGMA-EOP Working Group (site overview in **Table S1**). Of these, 57.6% of patients were classified as early-onset schizophrenia (EOS; n=223), 26.9% with affective psychoses (AFP; n=104), and 15.5% with other psychotic disorders (OTP; n=60). Details on site-wise recruitment procedures are presented in **Table S2**. Symptom severity was assessed with the Positive and Negative Syndrome Scale (PANSS) [48] or the Scale for the Assessment of Negative Symptoms (SANS) [49] and the Scale for the Assessment of Positive Symptoms (SAPS) [50]. Antipsychotic medication dose at the time of inclusion was converted to chlorpromazine-equivalent doses (CPZ) [51]. Demographic and clinical information and statistical tests of group differences for the mega-analysis dataset are presented in **Table 1**. Missing variables are reported in **Note S1**. Demographic and clinical information stratified by site are given in **Table S3**. One additional dataset (n=20 EOP; n=25 CTR) was included only in the supplementary meta-analysis and is described separately in **Table S4**.

**Table 1.**
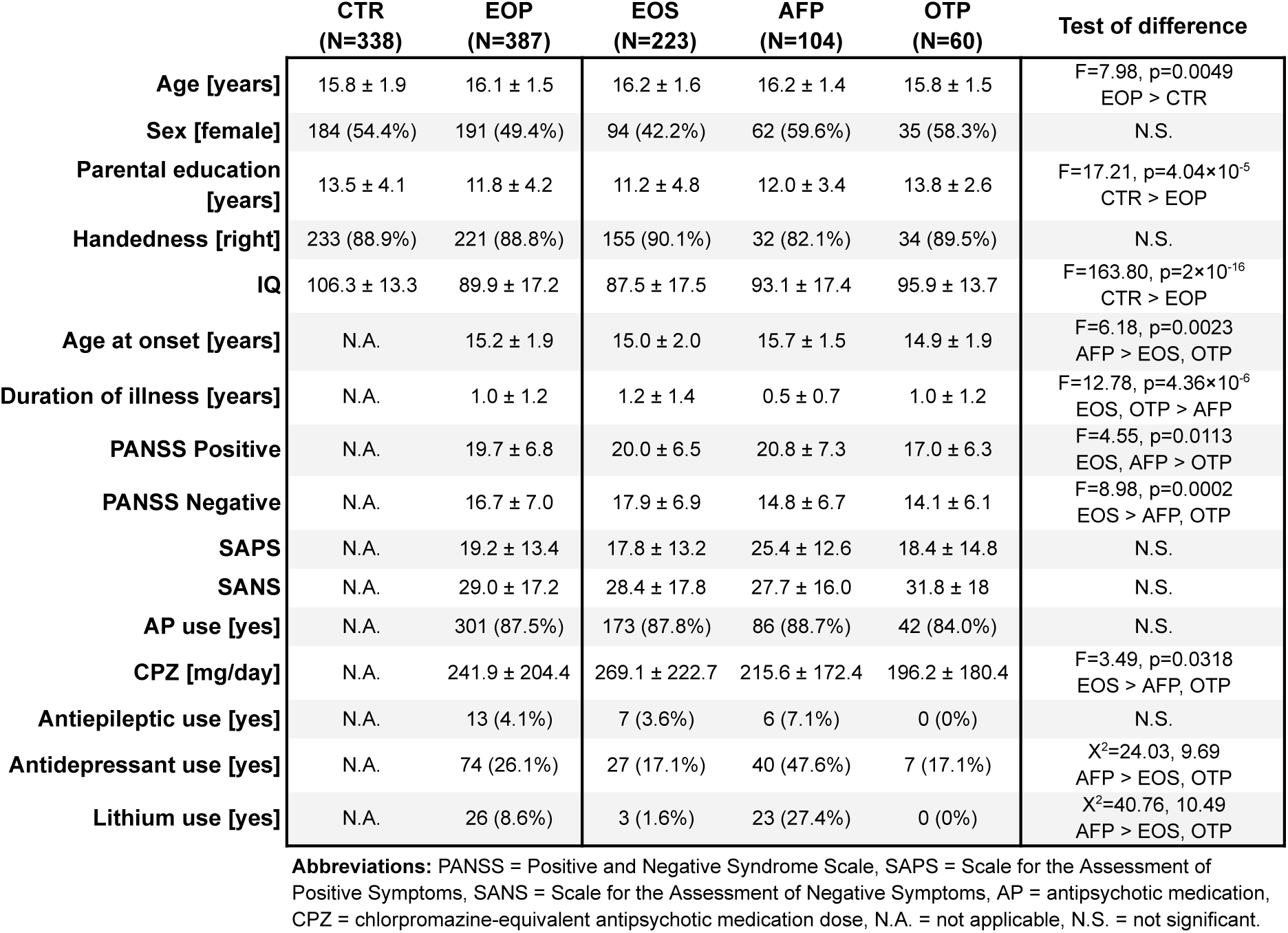
Demographic and clinical information. Sample description for the mega-analysis dataset. Continuous variables are given as means ± standard deviations and categorical variables as counts (percentage of non-missing). Group differences were assessed using Analysis of Variance (ANOVA) for continuous and *χ^2^*tests for categorical variables. Comparisons were made between EOP and CTR for demographics and IQ, and among EOS, AFP, and OTP for clinical variables.

### MRI acquisition and processing

Acquisition parameters for each scanner are detailed in **Table S5**. T1-weighted brain images were processed with the *recon-all* pipeline of FreeSurfer (v7.1.0 or above) [37,38] to extract mean cortical thickness, surface area, cortical volume, and LGI for the 68 regions of interest of the Desikan-Killiany atlas (34 per hemisphere). See **Figure S1** for an overview of this atlas. Briefly, the *recon-all* pipeline corrects for intensity nonuniformities, creates cortical surface meshes representing the cortex/non-brain and grey/white matter interfaces, and parcellates the cerebral cortex using sulcogyral folding patterns. Intracranial volume (ICV) was computed with the Sequence Adaptive Multimodal SEGmentation (SAMSEG) tool in FreeSurfer [52], which may be more robust compared to registration-based approaches [53]. We used a harmonized quality control procedure, including visual inspection of T1-weighted images and surface reconstructions (**Note S2**). To adjust for scanner-related technical variability, we used ComBat harmonization [54,55] as described in **Note S3**.

### Statistical analyses

Statistical analyses were conducted in R (version 4.2.4) [56]. For each cortical metric and set of analyses, we controlled the False Discovery Rate (FDR) with the Benjamini-Hochberg procedure [57]. Group differences in cortical metrics were considered statistically significant at FDR-adjusted p<0.05. Unless otherwise stated, only statistically significant results are reported. Cohen’s *d* effect sizes were computed from *t*-values using the *effectsize* package in R. Statistical model specifications and further details are given in **Note S4**.

#### Demographic and clinical group differences

We tested for group differences in age, sex, parental education, handedness, IQ, duration of illness, age at onset, symptom scores, antipsychotic medication use status (yes/no), CPZ-equivalent dose, and antiepileptic and antidepressant use status. Comparisons were performed for EOP and CTR for demographic variables and IQ, and between EOS, AFP, and OTP for clinical variables. Group differences in continuous variables were assessed using Analysis of Variance (ANOVA) with follow-up pairwise *t*-tests to determine the direction of differences if significant, and categorical variables were assessed with *χ^2^*tests.

#### Case-control differences

To test for case-control differences in cortical metrics, we fitted regression models adjusted for age, age^2^ and sex, with diagnostic group (CTR/EOP) as the variable of interest (main model). This model was fitted for each cortical metric and bilateral region, as well as for global cortical metrics, yielding 70 models for each cortical metric. We included age^2^ as a covariate given putative nonlinear aging effects in adolescence [58]. To quantify the hemispheric specificity of case-control differences, we computed the cortical asymmetry index, defined for each cortical region as the difference between measurements in the left and right hemispheres, divided by their mean [46]. We then fitted regression models with the asymmetry indices as outcomes with the same predictors as in the main model. To characterize between-site heterogeneity, we conducted meta-analyses with the *metafor* package in R [59] (**Note S6**), including an additional dataset (**Table S4**).

#### Diagnostic subgroup analyses

To assess case-control differences for diagnostic subgroups, we used the same model specification as in the main analysis, where the diagnostic group term replaced with diagnostic subgroup (EOS/AFP/OTP) with CTR as reference.

#### Regional specificity and adjustment for intracranial volume

To examine the regional specificity of case-control differences, we fitted regression models for each cortical region where the global values of each cortical metric were included as an additional covariate. These models assess regional group differences between EOP and CTR, adjusted for sex, age, and age^2^, relative to the global value of each cortical metric, and indicate case-control differences in addition to potentially broad diagnostic effects that are not region-specific. Since a prior ENIGMA-EOP study showed large case-control differences for ICV [60], we conducted an additional sensitivity analysis by fitting separate models adjusted for age, age^2^, sex, and ICV.

#### Medication effects and associations with clinical variables

Antipsychotic medication effects were assessed by contrasting patients currently using (n=301) with those not using (n=43) antipsychotic medication in separate regression models. Similarly, we contrasted patients using antidepressants (n=74) with non-users (n=208). We further assessed associations between regional cortical morphology and CPZ among patients currently using antipsychotic medication (n=268). We tested for associations between cortical morphology and PANSS negative and positive symptom scores, duration of illness, and age of onset by fitting separate regression models for each of these variables adjusted for age, age^2^, and sex. To quantify the relationship between continuous clinical variables and cortical metrics, we computed partial regression coefficients adjusting for sex, age, and age^2^ based on the fitted regression models with the *effectsize* package in R.

#### Interactions with age and sex and sex-stratified analyses

To test for interactions between diagnostic group and sex and age, we fitted two separate regression models including interaction terms: 1) age-by-group, adjusting for sex, and 2) sex-by-group, adjusting for age and age^2^. Furthermore, we performed complementary sex-stratified analyses by fitting regression models adjusted for age and age^2^, with diagnostic group (CTR/EOP) as the variable of interest for males and females separately.

#### Comparison to adults with schizophrenia and bipolar disorders

To compare case-control differences in EOP with those observed in adult SCZ and adult/youth BD, we first computed Pearson correlations based on Cohen’s *d* effect sizes from the case-control comparisons for EOP, EOS, AFP, and OTP, and the cortical alterations reported in two prior ENIGMA studies [39,40]. Differences between correlation coefficients were evaluated using Steiger’s test [61]. To assess group differences in the magnitude of effects, we conducted paired *t*-tests across regions and calculated the ratio of the mean regional effect sizes in EOP to that of each comparison group, yielding an index of the average proportional difference in effect size magnitudes across cortical regions.

Cohen’s *d* estimates for case-control differences in cortical thickness and surface area in adults with SCZ were obtained from the ENIGMA-SZ study of 4,474 adults with SCZ [39], with a sample-size weighted mean age of 32.3 years and range of 21.2-43.6 years. For BD, estimates were obtained from the ENIGMA-BD study of 1837 adults and 411 youths with BD (< 25 years at scanning) [40]. Youths with BD had a mean age of 21.1 years and a mean illness onset of 20.3 years. We extracted these effect sizes with the ENIGMA Toolbox (v2.01; https://github.com/MICA-MNI/ENIGMA) [62].

To ensure consistency in covariate adjustment and enable direct effect size comparison, we recomputed Cohen’s *d* for the case-control differences between EOP and healthy controls to use the same covariates as in these previous studies. Thus, in the comparison with SCZ, we computed effect sizes adjusted for sex and age but not age^2^. In the comparison with BD, we adjusted for sex and age for cortical thickness, and sex, age, and ICV for surface area. Finally, to statistically compare case-control differences for each region individually, we conducted *z*-tests following the approach utilized in a previous ENIGMA-EOP study [40]. See **Note S5** for further details on the statistical comparisons with prior ENIGMA studies.

## 3. Results

### Demographic and clinical group differences

EOP participants (mean age=16.1 years) were older than CTR (mean age=15.8 years), but sex distributions were similar. Parental education and IQ were lower in EOP than in CTR. We found an older age of onset and more antidepressant users in AFP relative to EOS and OTP. Duration of illness was higher in EOS and OTP relative to AFP. PANSS positive scores were higher in EOS and AFP relative to OTP, whereas PANSS negative scores were higher in EOS compared to both AFP and OTP. CPZ was higher in EOS compared to both AFP and OTP. Finally, there were more antidepressant and lithium users in AFP compared to EOS and OTP. Group comparisons for the other demographic and clinical variables were not statistically significant. See **Table 1** for further details on the group differences.

### Case-control differences

We observed a widespread pattern of lower thickness, area, volume and LGI in EOP relative to CTR. Cortical thinning was bilateral, with more regions showing lower thickness in the left than in the right hemisphere (left/right: 20/15 regions). In the right hemisphere, frontal regions were relatively preserved. The largest group differences were observed in the superior frontal region in both hemispheres (left/right: −0.36/-0.37). Area, volume, and LGI were lower across most cortical regions in EOP compared to CTR, with overall larger effect sizes than those for thickness. Global thickness was lower in EOP compared to CTR in both hemispheres (left/right: −0.36/-0.31). Similarly, global area (left/right: −0.42/-0.40), volume (left/right: −0.58/-0.56), and LGI (left/right: −0.39/-0.52) were lower in EOP compared to CTR. See **Figure 1a** for surface maps of effect sizes for significant differences in the case-control comparison and **Figures S2-S5** for bar plots of effect sizes for each cortical metric and hemisphere. See **Table S6** for model summaries for the analyses of global cortical metrics.

**Figure 1.**
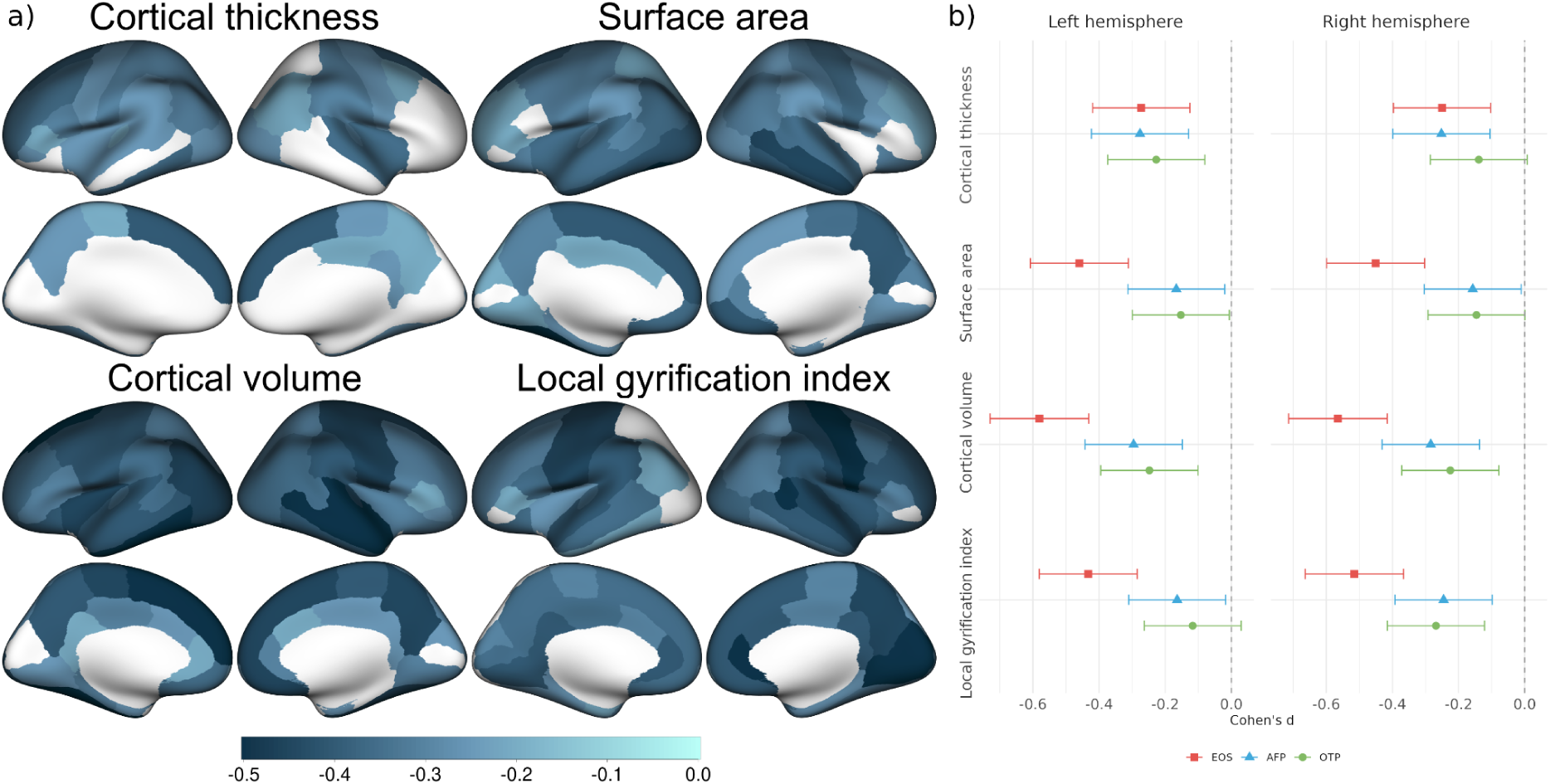
a) Cohen’s *d* effect sizes for cortical regions that differed significantly after FDR-correction between early-onset psychosis (EOP) and healthy controls, where darker colour indicates greater magnitude. b) Cohen’s *d* effect sizes for global cortical metrics stratified by the diagnostic subgroups, early-onset schizophrenia (EOS; *red rectangle*), affective psychosis (AFP; *blue triangle*), and other psychoses (OTP; *green circle*).

Cortical asymmetry analyses showed a leftward skew in EOP for thickness in the inferiortemporal (*d*=-0.26; *p_FDR_*=0.018) and superior parietal (*d*=-0.24; *p_FDR_*=0.027) regions and a rightward skew for LGI in the insula (*d*=0.22; *p_FDR_*=0.047) and the lateral occipital (*d*=0.23; *p_FDR_*=0.047) regions. See **Figure S6** for a caterpillar plot of asymmetry differences.

The meta-analysis indicated considerable heterogeneity between individual sites but the results supported the main findings. See **Figure S7** for forest plots with estimated group differences for each site and meta-analytic estimates for the global cortical metrics.

### Diagnostic subgroup analyses

Effect sizes for the global cortical metrics were similar across diagnostic subgroups for thickness, whereas alterations in EOS were greater for area, volume, and LGI (**Figure 1b**). We observed a fronto-temporal pattern of cortical thinning in EOS relative to CTR, whereas AFP and OTP showed a pattern of lower thickness in frontal and parietal regions. Area, volume, and LGI were consistently lower in EOS relative to CTR across the cerebral cortex. In contrast, regional surface area only differed significantly for a few cortical regions in AFP (5 regions) and OTP (3 regions) compared to CTR. In both AFP and OTP, lower LGI was mostly seen in the right hemisphere and the cingulate, precuneus, and cuneus for AFP. Only global thickness in the right hemisphere and global LGI of the left hemisphere in OTP did not differ relative to CTR. See **Figure 2** for surface maps with effect sizes for cortical regions that differed significantly in the diagnostic subgroup analyses. Thickness alterations in EOS had a greater correlation with AFP (*r*=0.78) than with OTP (*r*=0.53). Correlations between case-control alterations across diagnostic subgroups are given in **Table S7**.

**Figure 2.**
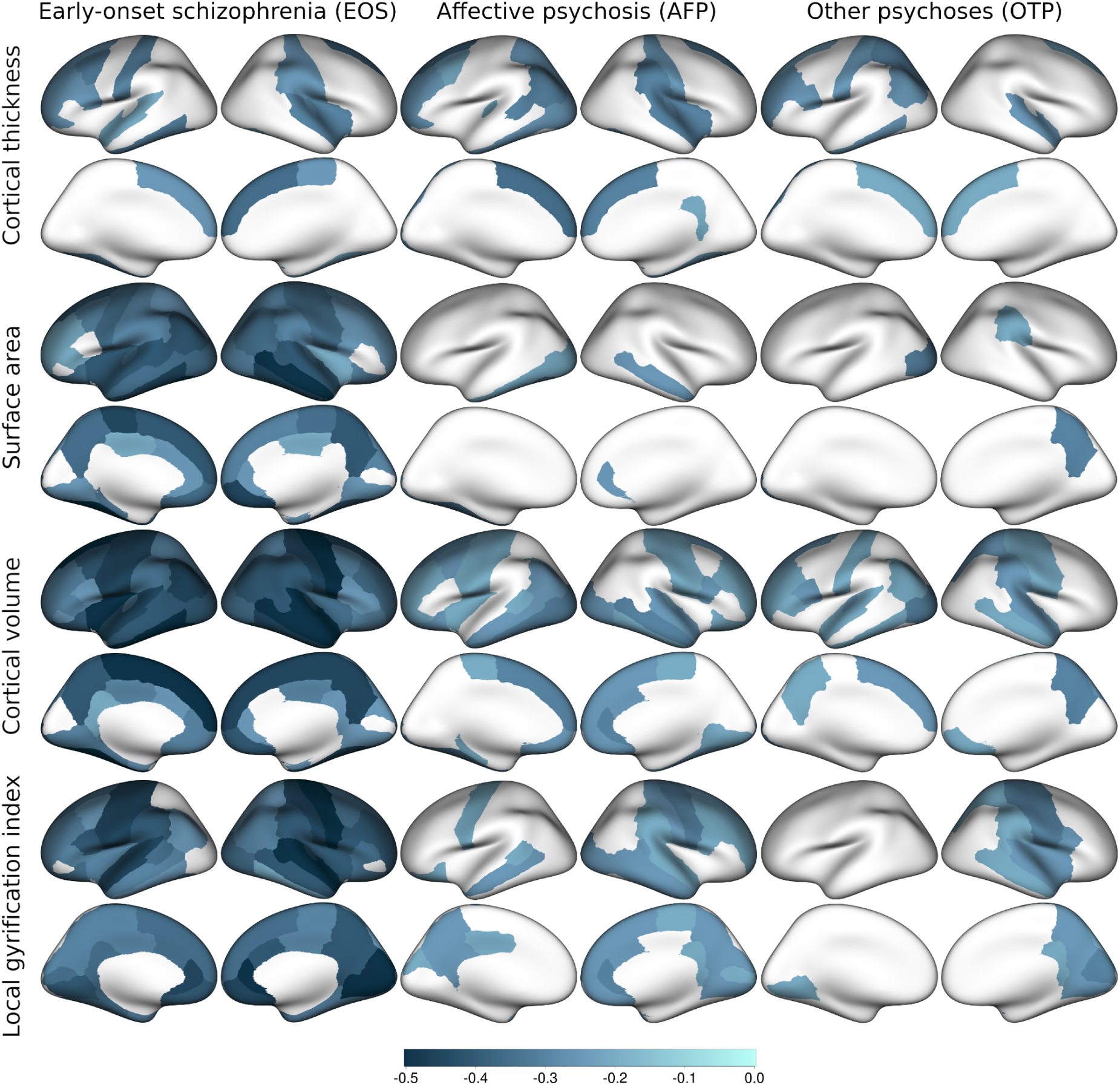
Cohen’s *d* effect sizes for cortical regions that differed significantly between early-onset schizophrenia (EOS), affective psychosis (AFP), other psychoses (OTP), and healthy controls. Columns correspond to diagnostic subgroups and rows depict statistically significant differences compared to healthy controls for cortical thickness, surface area, cortical volume and the Local Gyrification Index (LGI).

### Regional specificity and adjustment for intracranial volume

Adjusting for global thickness, we observed higher relative thickness in the cuneus in the left hemisphere (*d*=0.32) in EOP relative to CTR. Regional surface area was smaller in the fusiform gyrus in the left hemisphere (*d*=-0.28). No other case-control comparisons were statistically significant. See **Figures S8-S11** for bar plots of effect sizes for each cortical metric and hemisphere adjusted for global cortical metrics.

Adjusting for ICV, case-control differences were similar to those of the main analyses for thickness and LGI, whereas volume differences in the prefrontal cortex and the cingulate were largely explained by ICV. For area, only case-control differences in the inferior temporal lobe and in medial regions remained significant after ICV-correction. See **Figure S12** for surface maps with effect sizes for each cortical metric when adjusting for ICV.

### Medication effects and associations with clinical variables

Cortical metrics did not differ significantly between antipsychotic medication users compared to non-users and there were no significant associations with CPZ. See **Figure S13** for uncorrected partial correlations between CPZ and cortical metrics. We observed smaller area (*d*=-0.43; *p_FDR_*=0.034) and volume (*d*=-0.50; *p_FDR_*=0.003) in the right rostral anterior cingulate (rACC) in current antidepressant users compared to non-users among patients with EOP. For psychotic symptoms, only the association between the PANSS negative subscale and the volume of the left lingual region was statistically significant (partial *r*=-0.21; *p_FDR_*=0.014). See **Figures S14-S15** for partial correlations between the PANSS positive and negative subscales and cortical metrics. There were no associations between cortical morphology and duration of illness or age of onset.

### Interactions with age and sex and sex-stratified analyses

There were no significant interactions between diagnostic group and age or sex for any of cortical metric. In sex-stratified analyses, lower regional thickness was observed only in female patients with EOP compared to female controls. However, males with EOP showed global thinning in both hemispheres relative to male CTR. More regions showed smaller area in male than female patients compared to sex-matched CTR, while volume and LGI alterations were more similar across sexes. All global cortical metrics differed from those of CTR in both male and female patients (**Figure 3**). See **Figures S16-S17** for surface maps of regional effect sizes for significant regions, and **Figures S18-22** for bar plots of effect sizes across all cortical regions from the sex-stratified analyses.

**Figure 3.**
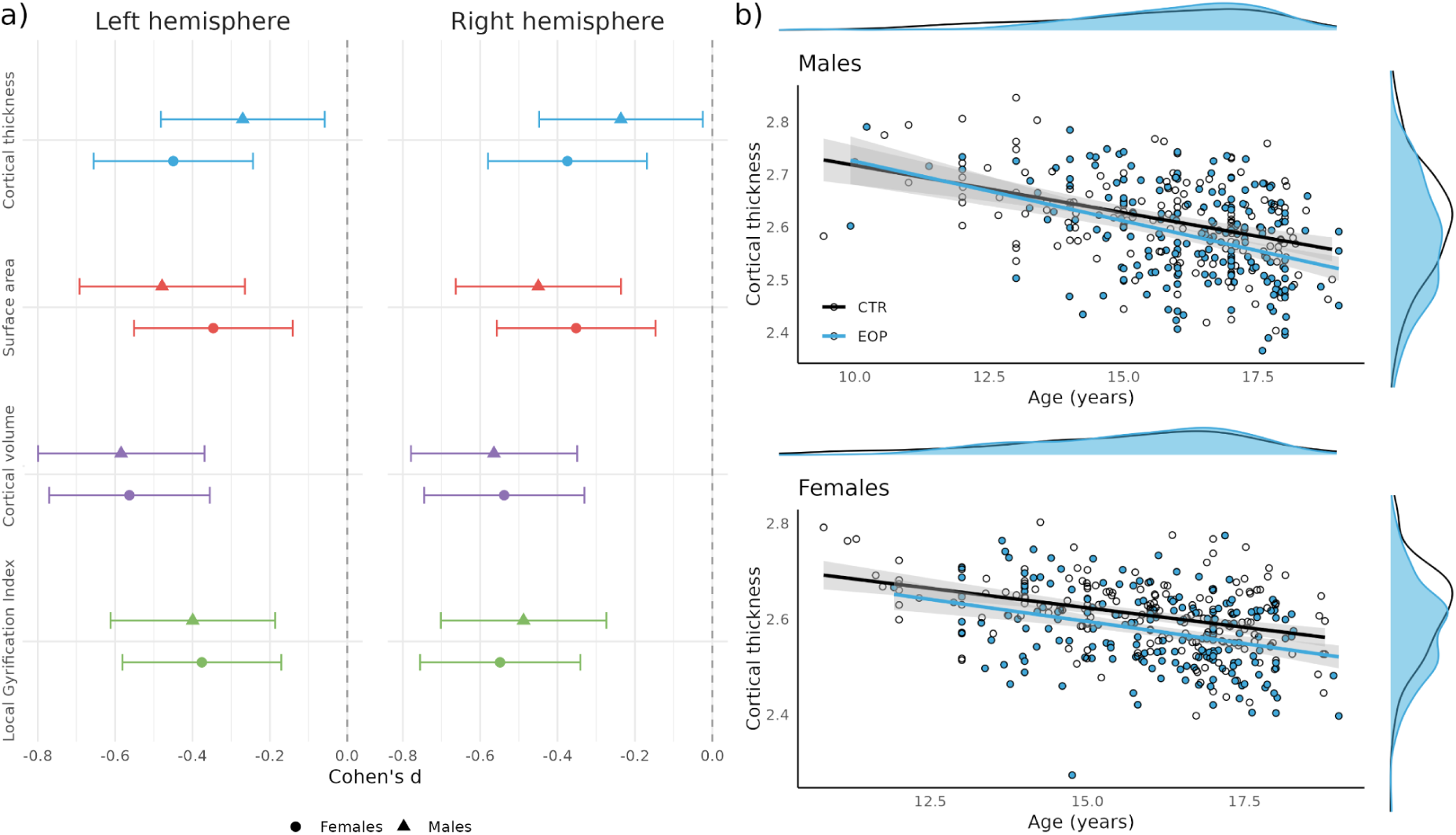
a) Cohen’s *d* effect sizes for females (*circles*) and males (*triangles*) for global cortical thickness, surface area, cortical volume, and Local Gyrification Index (LGI) for each hemisphere. The bars denote 95% confidence intervals. b) Marginal plots showing global cortical thickness as a function of age stratified by sex with early-onset psychosis (EOP) represented with blue and healthy controls with white circles. The blue distributions depict thickness and age distributions for EOP and the black outlines depict the same distributions for controls.

### Comparison to adults with schizophrenia and bipolar disorders

We observed strong correlations between regional Cohen’s *d* effect sizes for EOP and adult SCZ for thickness (*r*=0.62; *p*=1.4×10^-8^) and moderate correlations for area (*r*=0.49; *p*=2.0×10^-5^). EOP thickness alterations were more strongly correlated to those of adult BD (*r*=0.61; *p*=4.0×10^-8^) than those of youth BD (*r*=0.35; *p*=0.004). Surface area alterations in EOP did not correlate significantly with either BD group. In the diagnostic subgroup analyses, we observed strong correlations between cortical thickness alterations in EOS and AFP with those of adult SCZ and adult/youth BD, whereas the correlations for cortical thickness alterations in OTP were weak (**Figure 4b**).

**Figure 4.**
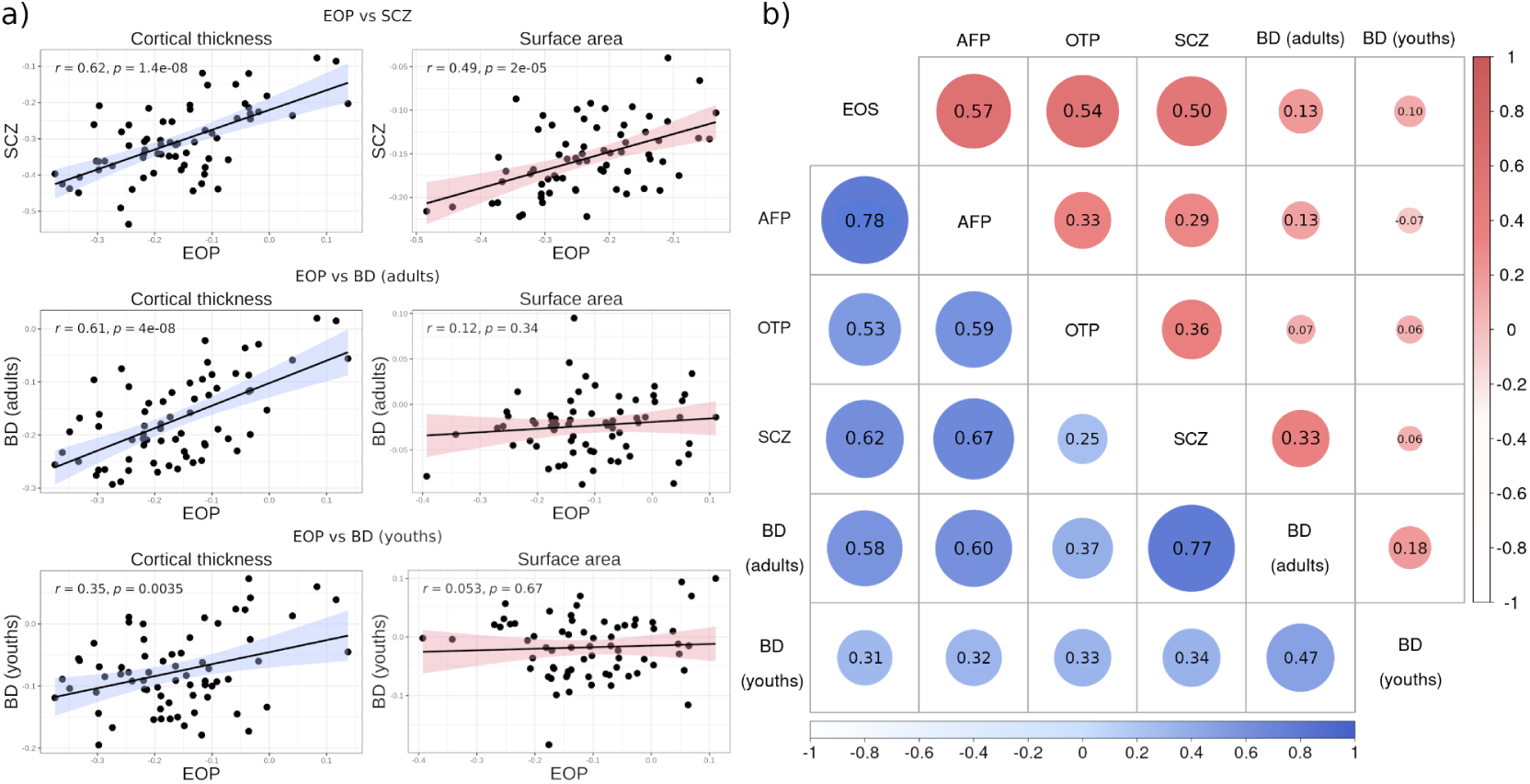
a) Correlations between case-control differences in cortical thickness (*blue*) and surface area (*red*) between early-onset psychosis (EOP), adults with schizophrenia (SCZ) and bipolar disorders (BD), as well as youths with BD. Regional Cohen’s *d* effect sizes for EOP are depicted on the *x*-axes and the *y*-axes represent effect sizes for each comparison group. b) Correlations between case-control differences for early-onset schizophrenia (EOS), affective psychosis (AFP), other psychoses (OTP), and adults with SCZ and BD, as well as youths with BD. The lower diagonal shows correlations for cortical thickness (*blue*) and the upper diagonal shows correlations for surface area (*red*). Darker colours and larger circles indicate stronger correlations.

The magnitude of Cohen’s *d* effect sizes for cortical thickness were significantly smaller in EOP compared to adult SCZ (*p*=2.2×10^-16^), but greater compared to youth BD (*p*=7.7×10^-9^). Effect sizes for cortical thickness in EOP did not differ significantly from those of adult BD. Post hoc analyses indicated that the average effect sizes for cortical thickness were 0.53 times those of adult SCZ and 2.11 times those of youth BD. For surface area, the magnitude of the effect sizes was greater in EOP compared to those of adult SCZ (*p*=4.5×10^-11^), adult BD (*p*=5.1×10^-9^), and youth BD (*p=*7.0×10^-9^). Post hoc analyses indicated that the effect sizes for surface area were 1.51 times those of adult SCZ, 4.62 times those of adult BD, and 6.11 times those of youth BD.

Regional effect size comparisons showed significantly lower effect sizes in EOP for thickness relative to adult SCZ across multiple regions, with more limited differences but greater effect sizes in EOP relative to both adults and youth BD. For area, effect sizes in EOP were greater than those of adult SCZ and adult/youth BD. See **Figures S22-S24** for bar plots of Cohen’s *d* effect sizes in EOP and each comparison group and **Tables S8-S14** for results from the comparisons of correlations and regional effects.

## 4. Discussion

In the largest study on cortical morphology in adolescent psychosis to date, we found widespread lower cortical thickness, surface area, cortical volume, and LGI in EOP relative to healthy controls. There were more pronounced deficits in surface area, cortical volume, and LGI in EOS than in AFP and OTP. The spatial pattern of cortical thickness alterations in EOP closely resembled those in adult SCZ and BD, whereas surface area alterations correlated only with those of adult SCZ. Moreover, the magnitude of cortical thickness deficits in EOP was approximately half that of adult SCZ, while surface area deficits were about 1.5 times greater in EOP. These findings suggest that cortical thickness alterations may reflect both static and progressive effects, whereas surface area alterations may indicate greater neurodevelopmental involvement in EOP compared to adult-onset SCZ.

A key finding of this study was the widespread nature of the case-control differences across cortical metrics, suggesting a broad pattern of diffuse alterations. This was supported by the regional specificity analyses where only two regions differed significantly relative to controls when adjusting for global case-control deficits. Similarly, the cortical asymmetry analyses only showed different lateralization patterns in EOP for three regions relative to healthy controls, indicating that case-control differences in cortical morphology were largely bilateral. The spatial distribution of case-control differences was greatest for cortical volume, LGI and surface area, with fewer significant differences for cortical thickness. Notably, no cortical thickness alterations were observed in the cuneus, lingual, fusiform, and middle temporal regions in either hemisphere, and the right frontal regions were relatively preserved. The broad pattern of cortical alterations is in line with a prior ENIGMA-EOP study on VBM by Si et al. (2024), but differed from some smaller studies reporting more focal alterations in adolescent psychosis [63,64]. The greater extent of case-control differences in the present study, compared to some prior studies, likely reflects the high statistical power, enabling the detection of more subtle brain structural deviations.

EOS showed more extensive and greater regional differences in surface area, cortical volume, and LGI compared to AFP and OTP. In contrast, global thickness differences were similar across subgroups, although AFP exhibited more significant regional alterations, particularly in the left parietal and occipital lobes. Cortical thickness alterations were highly correlated between EOS and AFP (*r*=0.78) and moderately between EOS and OTP (*r*=0.53). As cortical thickness deficits may reflect both static and progressive processes, more pronounced subgroup differences may arise later in the illness course. The findings partly support the hypothesis that neuroanatomical alterations are more pronounced in EOS. However, subgroup sizes differed (57.6% EOS; 26.9% AFP; 15.5% OTP), which likely influenced statistical sensitivity. This highlights the importance of large samples in diagnostic subgroup analyses, particularly given the marked clinical heterogeneity in EOP [65]. Diagnostic assessment of adolescents poses additional challenges, as most criteria are validated in adults and often depend on time-based criteria that are difficult to assess in adolescents. Diagnostic stability is especially low in OTP compared to EOS or AFP [66,67], warranting cautious interpretation of findings in this subgroup.

Longitudinal studies have reported accelerated cortical GM loss, notably in the frontal lobe, in EOP compared to healthy controls [25–27,68,69]. Such progressive cortical alterations could explain the preservation of right frontal cortical thickness seen in EOP and smaller effect sizes relative to adults with SCZ. If cortical thinning is progressive, alterations in adolescents with psychosis may eventually resemble the patterns seen in adults, a process hypothesized to reflect accelerated synaptic pruning during adolescence [70,71]. Furthermore, brain connectivity has been proposed to shape the spatial distribution of cortical thinning in SCZ, with mutually atrophic effects driving coordinated thinning across distant regions [72], supported by studies linking cortical thinning to functional and structural cortico-cortical connectivity [73–77]. However, the role of connectivity in driving these alterations remains unclear, as are the underlying neurobiological mechanisms, which may be confounded by antipsychotic use, lifestyle, adversity, and other exogenous factors. Future research should study the progressive aspects of brain structure in EOP, how brain connectivity relates to cortical alterations, and whether these changes reflect psychosis-specific effects or broader neurodevelopmental vulnerabilities [78–80].

Compared to previous large-scale ENIGMA studies [39,40], we found that surface area alterations in EOP correlated significantly with those of adult SCZ, but not BD, with the strongest correlations for EOP (*r*=0.49) and EOS (*r*=0.50). Importantly, we observed greater surface area alterations in EOP than in adult SCZ. While cortical thinning may be, at least partly, progressive, surface area and LGI stabilize in early adolescence and are considered more closely linked to neurodevelopmental processes [60,81,82]. These findings align with the ENIGMA-EOP study by Gurholt et al. (2022), where the largest case-control effect size was reported for ICV [60], which was greater (*d*=-0.39) than for adult SCZ (*d*=-0.12) [83]. In the present study, case-control differences in surface area were partly explained by ICV, and only one region, the left fusiform area, remained significant after adjusting for global surface area. This suggests that surface area alterations are global, may reflect pre-existing brain structural differences, are more specific to psychotic disorders, and may indicate a greater neurodevelopmental burden in EOP than in adult-onset SCZ.

Although sex-by-group interactions were not significant, stratified analyses revealed significant regional cortical thinning only in female patients with EOP relative to same-sex healthy controls. This contrasts with findings from the ENIGMA-EOP study by Barth et al. (2023), where male patients exhibited greater abnormalities of white matter microstructure than female patients [84]. Nevertheless, both sexes showed significant global cortical thinning relative to controls. Thus, cortical thinning in male EOP was less pronounced than in female EOP relative to their same-sex counterparts. Adolescence is a critical period of brain maturation, marked by substantial sex differences [85–87]. If cortical thinning reflects progressive pathology, earlier maturation in female patients may be linked to earlier and more pronounced cortical thinning. Over time, both sexes may converge toward the cortical thinning pattern seen in adult SCZ [72]. Future studies should further investigate sex-specific mechanisms in EOP, ideally incorporating hormonal and pubertal markers.

We found no significant associations between cortical metrics and PANSS positive scores, antipsychotic use or dose, or age-by-group interactions. However, negative PANSS scores were associated with lower cortical volume in the left lingual region. Additionally, antidepressant users showed significantly lower surface area and volume in the right rACC compared to non-users. The rACC has been implicated in depression and treatment response in Major Depressive Disorder [88–90], thus this finding may reflect comorbid depressive symptoms, which are common in EOP [91]. In contrast to Si et al. (2024), we found no association between cortical volume and antipsychotic dose or age of onset. Given the strong correlation between age of onset and age (*r*=0.74), and potential site-specific recruitment biases, disentangling these effects is challenging. Similarly, illness onset and cumulative medication exposure are closely linked, complicating causal inference. Longitudinal studies, especially in high-risk individuals, are needed to clarify how cortical morphology relates to medication exposure, illness duration, and age of onset.

Some important limitations apply to the present study. The study design does not permit causal inference, notably regarding medication effects and psychotic symptom severity, which vary over time. We lacked information on pubertal staging, which should be considered in future studies on adolescents. The data was collected from multiple research sites and MRI scanners, introducing both clinical and technical variability. While this may have introduced noise, it also provided an opportunity to assess between-site variability across diverse clinical settings via meta-analysis. In line with current recommendations, we used the scanner harmonization method, ComBat, which has been shown to attenuate scanner-related differences and increase statistical power, but residual effects of scanner differences may still have affected the results. Similarly, variations in inclusion/exclusion criteria across sites can lead to differences in diagnostic distributions, age ranges, or illness duration and severity. Such confounding between clinical characteristics and site is difficult to address statistically and represents an important limitation of multi-site studies.

### Conclusions

In the largest study of cortical morphology in adolescents with EOP to date, we found widespread lower cortical thickness, surface area, cortical volume, and LGI. Alterations were most pronounced in EOS and were independent of medication use, illness duration, or symptom severity, except for limited associations with negative symptoms and antidepressant use. The pattern of cortical thickness alterations closely resembled that seen in adults with SCZ and BD. In contrast, the pattern of surface area alterations only correlated with those of adults with SCZ, where the magnitude of the effects was, on average, 1.5 times in EOP than those of adults with SCZ. The findings suggest that greater surface area alterations may serve as a neurobiological signature of early-onset psychosis, and the larger effect sizes for surface area and LGI in adolescent psychosis highlight the potential impact of aberrant neurodevelopment on cortical morphology in this clinical group.

## Data Availability

Data analyzed in the present study may be accessed upon reasonable request to the authors and the ENIGMA-EOP Working Group.

## Acknowledgements

We are grateful to the clinical personnel and participants at contributing sites who made this research possible. We are also thankful for the contributions of Prof. Gianfranco Spalletta, who sadly passed away before the completion of this manuscript. This study made use of computing facilities provided by the Services for Sensitive Data (TSD), IT-department (USIT), University of Oslo, Norway. The following research grants facilitated the study: ENIGMA - US National Institutes of Health (NIH): R01-EB015611, R01-MH116147 (Sex Differences, to PMT), R01-MH117601, S10-OD023696, R01-MH129742 (ENIGMA-Bipolar, to PMT), R01-MH134004. The ENIGMA Working Group acknowledges the NIH Big Data to Knowledge (BD2K) award for foundational support and consortium development (U54 EB020403 to PMT). For a complete list of ENIGMA-related grant support please see here: http://enigma.ini.usc.edu/about-2/funding/. BARCELONA - Spanish Ministry of Health, Instituto de Salud Carlos III "Health Research Fund"/FEDER funds: PI17/00741; PI18/00976, PI21/00519, PI20/00654, PI21/00330, FORT23/00002_SUGR_G6, PI24/00279; PI24/01052; Brain and Behaviour Research Foundation: 26731; Department of Health, Government of Catalonia: SLT006/17/00362; Marato TV3: 202232-30-31 and 202210-10; Alicia Koplowtiz Foundation; Pons Bartran Legacy: FCRB-IPB2-2023; Maria and Nuria Cunillera Legacies. MADRID - Spanish Ministry of Science and Innovation, Instituto de Salud Carlos III (ISCIII), co-financed by the European Union, ERDF Funds from the European Commission, "A way of making Europe": Redes Temáticas-ISCIII G03-032; RETICS (REM-TAP Network) RD06/0011, PS09/01442, PI12/1303, PI22/01824, PI22/01621, PI23/00625; Consorcio Centro Investigación Biomédica en Red (CIBER): CB/07/09/0023; European Union: FP7-HEALTH-2013-2.2.1-2-603196 (Project PSYSCAN), METSY: FP7-HEALTH-2013-2.2.1-2-602478 (Project METSY); Madrid Regional Government: S2010/BMD-2422, B2017/BMD-3740, S2022/BMD-7216 (AGES 3-CM); Fundación Familia Alonso: FIBHGM-CCA004-2018; Fundación Alicia Koplowitz: FAK13. ROME - Italian Ministry of Health: Ricerca Corrente 25. OXFORD - MRC G0500092. KCL-1 - Brain & Behavior Research Foundation (prev. NARSAD). KCL-2 - Dutch Research Council (NWO): VIDI 0452-07-007 (to LK). UCLA - National Institute of Mental Health: P50MH066286, U01MH081902. SCAPS - Swedish Research Council: 521-2014-3487, 2017-00949; Formas: 259-2012-31. UIO-PSI - South-Eastern Norway Regional Health Authority: 2004-259, 2006-186. YTOP: South-Eastern Norway Regional Health Authority: 2017-097, 2017-112, 2019-104, 2020-020, 2021-070, 2022-080, 2024-012; Research Council of Norway: 223273, 274359, 2137000, 250358, 288083, 323951; K. G. Jebsen Foundation: SKGJ-MED-008.

