## Supplementary Materials for "Cerebral cortical alterations in adolescent early-onset psychosis: a surface-based morphometry mega-analysis"

#### Table of contents


##### 1. Supplementary Notes

**Note S1.** Further information on clinical assessments.
**Note S2.** Quality control procedure.
**Note S3.** Scanner harmonization.
**Note S4.** Regression model specifications.
**Note S5.** Comparison to prior ENIGMA studies.
**Note S6.** Meta-analysis across sites.

##### 2. Supplementary Figures

**Figure S1.** Overview of cortical regions in the Desikan-Killiany atlas.
**Figure S2-S5.** Main analysis - Bar plots of effect sizes.
**Figure S6.** Asymmetry analyses - Caterpillar plot of asymmetry indices.
**Figure S7.** Meta-analysis - Forest plots for global cortical metrics.
**Figure S8-S11.** Regional specificity analyses - Bar plots of effect sizes.
**Figure S12.** ICV-adjusted analyses - Surface maps of effect sizes.
**Figure S13.** CPZ - Surface maps with uncorrected partial correlations.
**Figure S14.** PANSS positive - Surface maps with uncorrected partial correlations.
**Figure S15.** PANSS negative - Surface maps with uncorrected partial correlations.
**Figure S16-S21.** Sex-stratified analyses - Surface maps and bar plots stratified by sex.
**Figure S22-S24.** Effect size comparison with previous ENIGMA studies.

##### 3. Supplementary Tables

**Table S1.** Overview of participating research sites.
**Table S2.** Recruitment procedures and criteria for each site.
**Table S3.** Site-wise demographic and clinical information.
**Table S4.** Demographic and clinical information of additional dataset.
**Table S5.** MRI acquisition parameters for each site.
**Table S6.** Model summaries for global cortical metrics.
**Table S7.** Correlations between case-control alterations across diagnostic subgroups.
**Table S8-S14.** Comparisons of correlations and effect sizes.

##### 4. References

### 1. Supplementary Notes

#### Note S1 - Further information on clinical assessments

Positive and Negative Syndrome Scale (PANSS) [1] symptom scores were available for 306 participants, while the Scale for the Assessment of Negative Symptoms (SANS) [2] and the Scale for the Assessment of Positive Symptoms (SAPS) [3] were available for 42 participants. Therefore the symptom severity analyses were conducted for the PANSS. Age of onset and duration of illness were missing for 26 participants. Information on current antipsychotic medication use status (yes/no) was available for 344 participants, of which 301 (87.5%) used antipsychotic medication at inclusion. Chlorpromazine-equivalent doses (CPZ) [4] of antipsychotic medication at inclusion were available for 306 participants.

#### Note S2 - Quality control procedure

Quality control (QC) was conducted using a dual approach: 1) We identified outliers using the Median Absolute Deviation (MAD) approach [5] implemented in the *routliers* package in R, with a MAD threshold of 4. The MAD is less biased by sample size and the presence of extreme outliers compared to the mean. Outliers flagged for any cortical metric and region were inspected visually to ensure that the surface reconstruction was acceptable. In the event of major errors or poor image quality, the participant was flagged and excluded from all further analyses. 2) We created QC images for each participant comprising coronal and axial views of the original T1-weighted images with Sequence Adaptive Multimodal SEGmentation (SAMSEG) [6] segmentations overlain. These were inspected visually for all participants, and if major segmentation errors were identified, or image quality was deemed to be unacceptable, the participant was excluded from further analyses. In the dataset used in the mega-analyses, we excluded 27 participants due to poor image quality, 2 participants due to processing errors, and 13 participants due to image artifacts, usually related to dental braces or movement in the scanner.

#### Note S3 - Scanner harmonization

To adjust for scanner-related variation, we applied ComBat harmonization for each cortical metric, i.e., cortical thickness, surface area, cortical volume, and the Local Gyrification Index (LGI), as well as segmentation-based total intracranial volume (ICV), separately. ComBat was developed to adjust for batch effects in microarray experiments and has become widely used for adjusting for scanner-related variation in neuroimaging [7–9]. We used empirical Bayes to leverage within-metric variation across all cortical regions of the Desikan-Killiany atlas [10] and specified age, sex, and case-control status as biological variables of interest.

**Note S4 - Regression model specifications**

Regression models were fitted with the *lm* command from the *stats* package in R [11]. To control the False Discovery Rate (FDR), we used the *p.adjust* command from the same package. FDR-correction was performed for each cortical metric separately, e.g., for the main analysis of cortical thickness, we performed a total of 70 statistical tests (68 cortical regions + 2 global hemispheric thickness values). Cohen's *d* effect sizes were computed from *t*-values from the fitted regression models using the *effectsize* package in R.

In the following regression model specifications, $DV_{ij}$ represents the dependent variable, i.e., cortical thickness, surface area, cortical volume, or the Local Gyrification Index (LGI) for the *j*-th cortical region (one of the 34 regions per hemisphere) for the *i*-ith participant. Similarly, the predictors of the models are indexed by participant. The reference group was healthy controls for Dx (levels: EOP/CTR) and FullDx (levels: EOS/AFP/OTP/CTR). The variable of interest for each model is denoted in bold type.

Case-control differences

All participants: $DV_{ij} \sim Age_i + Age_i^2 + Sex_i + \mathbf{Dx_i}$

Diagnostic subgroup analyses

All participants: $DV_{ij} \sim Age_i + Age_i^2 + Sex_i + \mathbf{FullDx_i}$

Regional specificity and adjustment for intracranial volume

1. All participants: $DV_{ij} \sim Age_i + Age_i^2 + Sex_i + GlobalMetric_i + \mathbf{Dx_i}$
2. All participants: $DV_{ij} \sim Age_i + Age_i^2 + Sex_i + ICV_i + \mathbf{Dx_i}$

Medication effects and associations with clinical variables

1. Non-missing patients: $DV_{ij} \sim Age_i + Age_i^2 + Sex_i + \mathbf{AP_i}$
2. Non-missing patients: $DV_{ij} \sim Age_i + Age_i^2 + Sex_i + \mathbf{AD_i}$
3. Non-missing patients: $DV_{ij} \sim Age_i + Age_i^2 + Sex_i + \mathbf{CPZ_i}$
4. Non-missing patients: $DV_{ij} \sim Age_i + Age_i^2 + Sex_i + \mathbf{PANSS\text{-}POS_i}$
5. Non-missing patients: $DV_{ij} \sim Age_i + Age_i^2 + Sex_i + \mathbf{PANSS\text{-}NEG_i}$
6. Non-missing patients: $DV_{ij} \sim Age_i + Age_i^2 + Sex_i + \mathbf{Duration\ of\ illness_i}$
7. Non-missing patients: $DV_{ij} \sim Age_i + Age_i^2 + Sex_i + \mathbf{Age\ of\ onset_i}$

Interactions with age and sex and sex-stratified analyses

1. All participants: $DV_{ij} \sim Age_i + Sex_i + Dx_i + \mathbf{Age_i \times Dx_i}$
2. All participants: $DV_{ij} \sim Age_i + Age_i^2 + Sex_i + Dx_i + \mathbf{Sex_i \times Dx_i}$
3. Male participants: $DV_{ij} \sim Age_i + Age_i^2 + \mathbf{Dx_i}$
4. Female participants: $DV_{ij} \sim Age_i + Age_i^2 + \mathbf{Dx_i}$

**Note S5 - Comparison to prior ENIGMA studies**

To harmonize covariate adjustment across case-control comparisons, we recomputed regional effect sizes for EOP adjusted only for age and sex for cortical thickness, and similarly for surface area in the comparison with adult SCZ. For surface area comparisons involving adults and youths with BD, we also adjusted for ICV. This ensured consistency in the selection of covariates across group comparisons and enabled direct comparison of effect sizes. Pearson correlation coefficients were compared using Steiger's test [12].

We used paired *t*-tests to compare Cohen's *d* effect sizes in EOP with those in adult SCZ, adult BD, and youth BD. To further characterize the average proportional difference in effect sizes between groups, we calculated the ratio of the mean Cohen's *d* effect size in EOP to that of each comparison group. This provided a descriptive measure of the average proportional difference in effect size magnitudes across regions.

Finally, we performed additional pairwise comparisons between region-wise Cohen's *d* effect sizes using a similar approach as in a prior ENIGMA-EOP study [13]. Specifically, we performed *z*-tests based on the following formulas:

1) $$Z_{Diff} = \frac{d_{Group1} - d_{Group2}}{\sqrt{SE(d_{Group1})^2 + SE(d_{Group2})^2}}$$

2) $$p = 2\left[1 - \left(\Phi(\left|Z_{Diff}\right|)\right)\right]$$

In equation 1), $d_{Group1}$ is the estimated Cohen's *d* effect sizes for the reference group, i.e., EOP, and $d_{Group2}$ is that of the comparison group (adult SCZ, adult BD, or youths with BD), and SE is the standard error of the estimated Cohen's *d*. In equation 2) the p-value is computed using a two-tailed test where Φ is the standard normal cumulative distribution.

**Note S6 - Meta-analysis across sites**

For the meta-analysis, we first fitted separate linear regression models for each individual research site, adjusted for age, age$^2$, and sex, with diagnostic status as the main variable of interest. The models were fitted separately for global cortical thickness, surface area, cortical volume, and the Local Gyrification Index (LGI) in the right and left hemispheres. For sites with data acquired on multiple MRI scanners, we included scanner as a covariate. We then extracted the t-statistics associated with the Dx term, which were converted to Hedges' *g* with the *effectsize* package in R. The Hedges' *g* was used to correct for small-sample bias. We estimated the standard error, $SE_g$, of the Hedges' *g* with the formula:

$$SE_g = \sqrt{\frac{n_{EOP} + n_{CTR}}{n_{EOP} \cdot n_{CTR}} + \frac{g^2}{2 \cdot (n_{EOP} + n_{CTR})}}$$

The meta-analysis was conducted using a random-effects model with restricted maximum likelihood (REML), implemented in the *rma* function in the *metafor* package in R [14]. This model accounts for both within-study variance and between-study heterogeneity. Forest plots are shown in **Figure S7**, displaying site-specific Hedges' *g* effect sizes with 95% confidence intervals, alongside the pooled meta-analytic effect across research sites. The heterogeneity between studies, i.e., the proportion of variability explained by between-site heterogeneity rather than chance, was estimated as $I^2$ [15].

### 2. Supplementary Figures

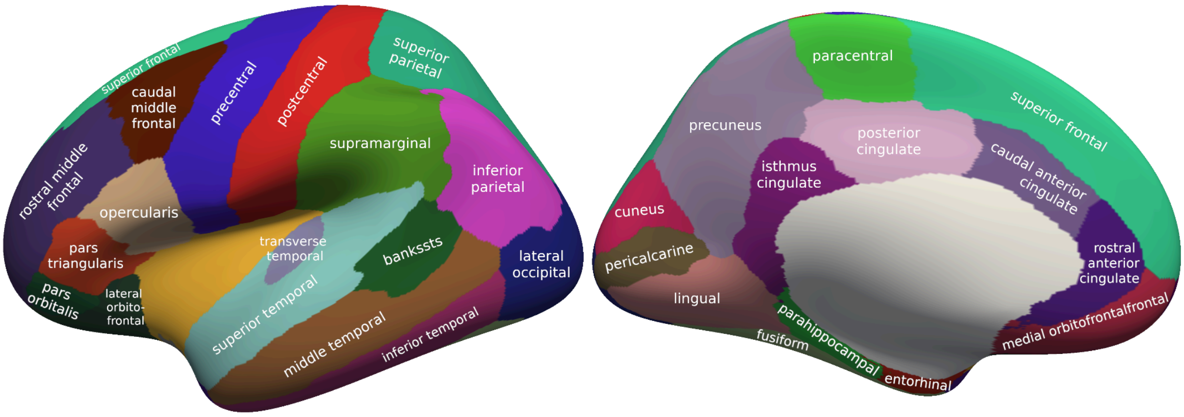


**Figure S1. Overview of cortical regions in the Desikan-Killiany atlas.** There are 34 regions per hemisphere with locations determined by the sulcogyral folding pattern [10].

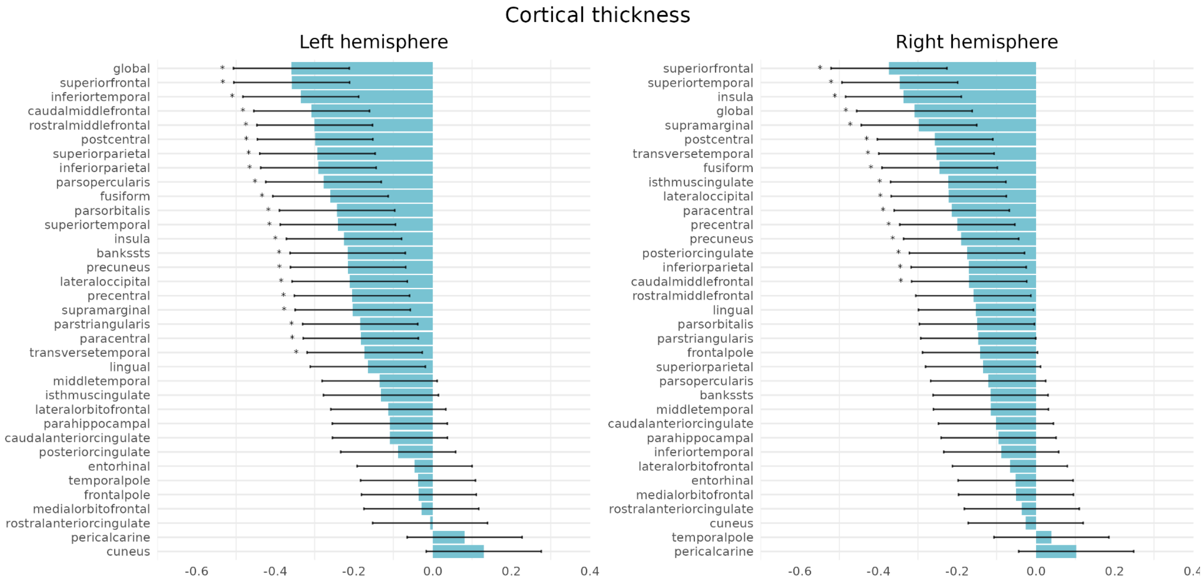


**Figure S2. Main analysis - Cortical thickness.** Cohen's *d* effect sizes for comparisons between patients with early-onset psychosis (EOP) and healthy controls from models adjusted for sex, age, and age$^2$. Effect sizes are ranked by magnitude; significant differences after correction for multiple testing are marked with asterisks.

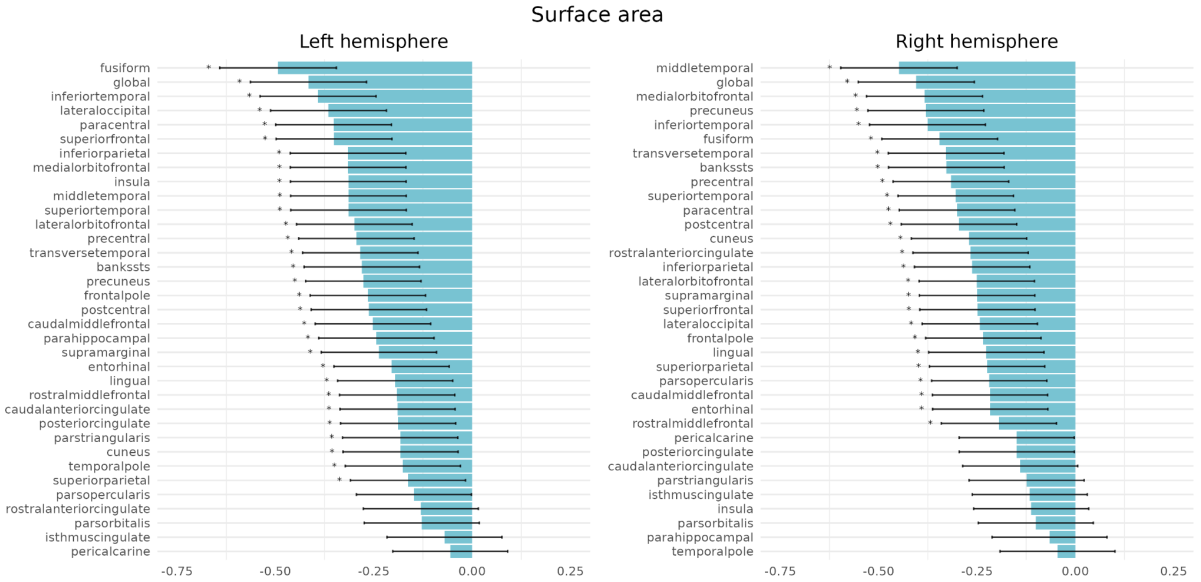


**Figure S3. Main analysis - Surface area.** Cohen's *d* effect sizes for comparisons between patients with early-onset psychosis (EOP) and healthy controls from models adjusted for sex, age, and age$^2$. Effect sizes are ranked by magnitude; significant differences after correction for multiple testing are marked with asterisks.

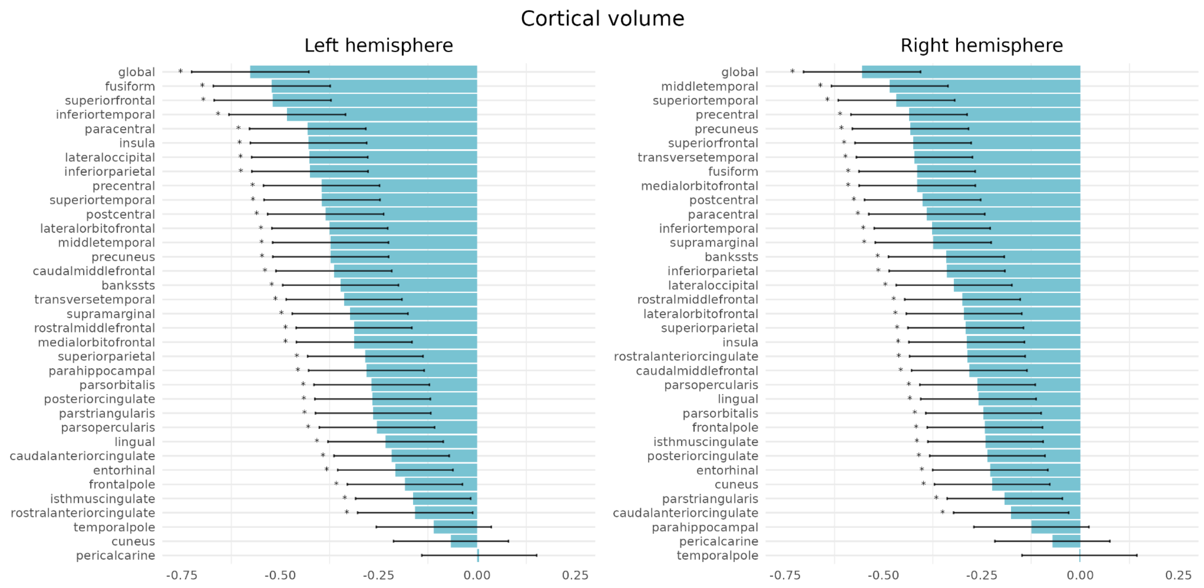


**Figure S4. Main analysis - Cortical volume.** Cohen's *d* effect sizes for comparisons between patients with early-onset psychosis (EOP) and healthy controls from models adjusted for sex, age, and age$^2$. Effect sizes are ranked by magnitude; significant differences after correction for multiple testing are marked with asterisks.

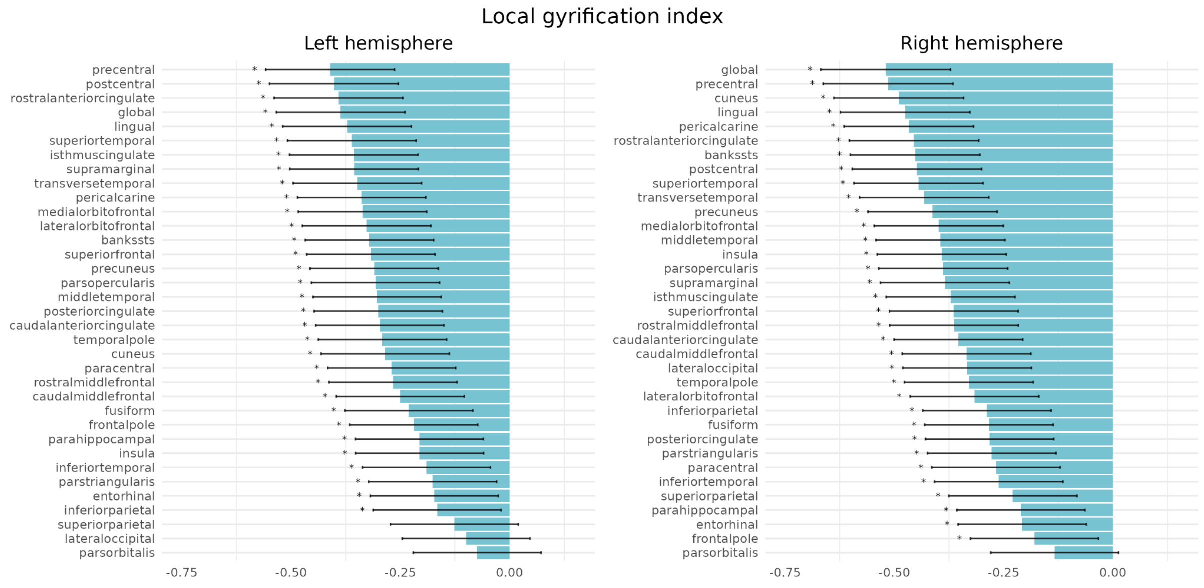


**Figure S5. Main analysis - Local Gyrification Index (LGI).** Cohen's *d* effect sizes for comparisons between patients with early-onset psychosis (EOP) and healthy controls from models adjusted for sex, age, and age$^2$. Effect sizes are ranked by magnitude; significant differences after correction for multiple testing are marked with asterisks.

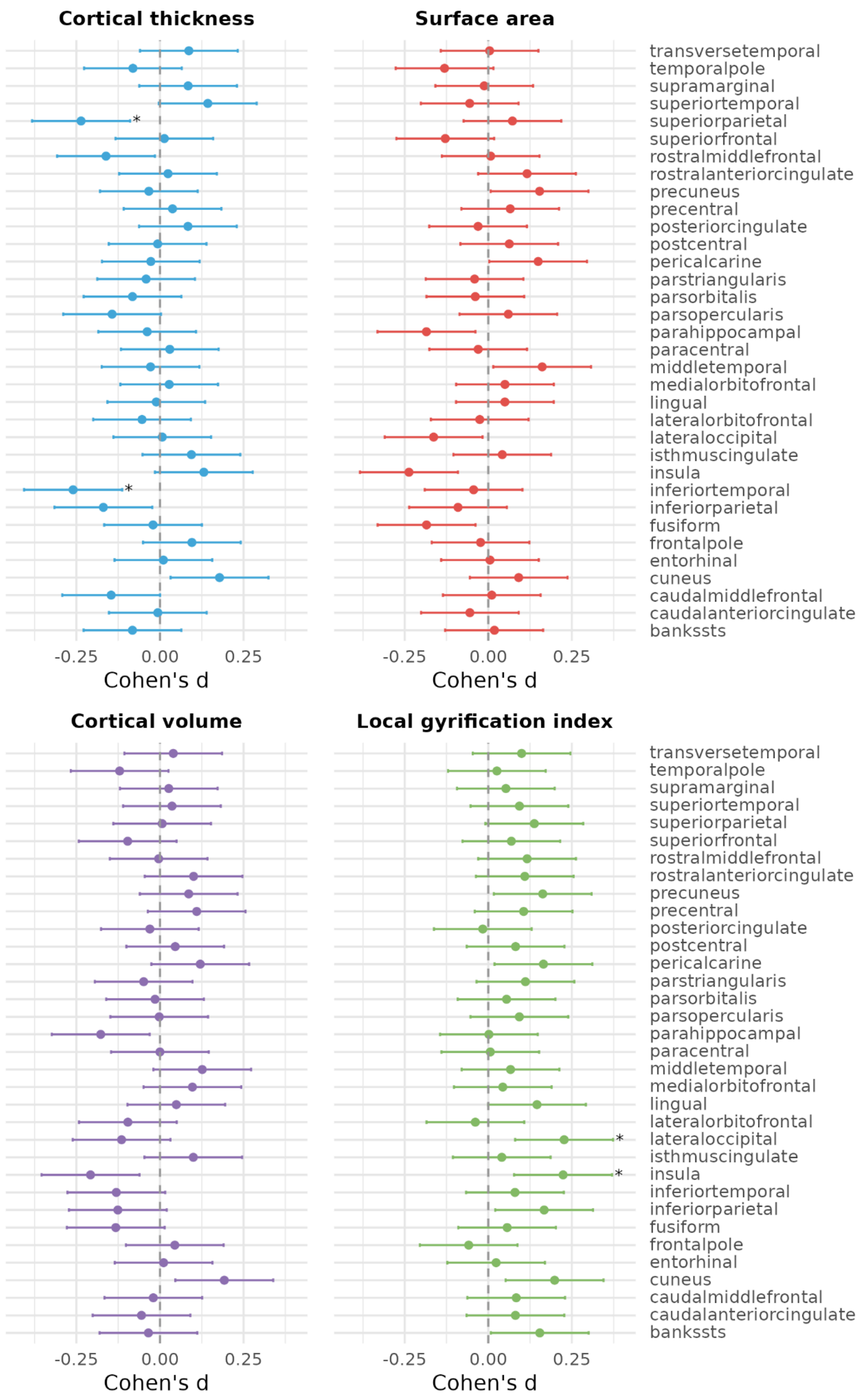


**Figure S6. Cortical asymmetry analyses.** The vertical bars depict Cohen's *d* effect sizes and their 95% confidence intervals for case-control comparisons in early-onset psychosis (EOP) vs healthy controls for the asymmetry index, i.e., left - right hemisphere divided by the mean, for each cortical region. Asterisks depict differences between EOP and healthy controls that were statistically significant after correction for multiple testing.

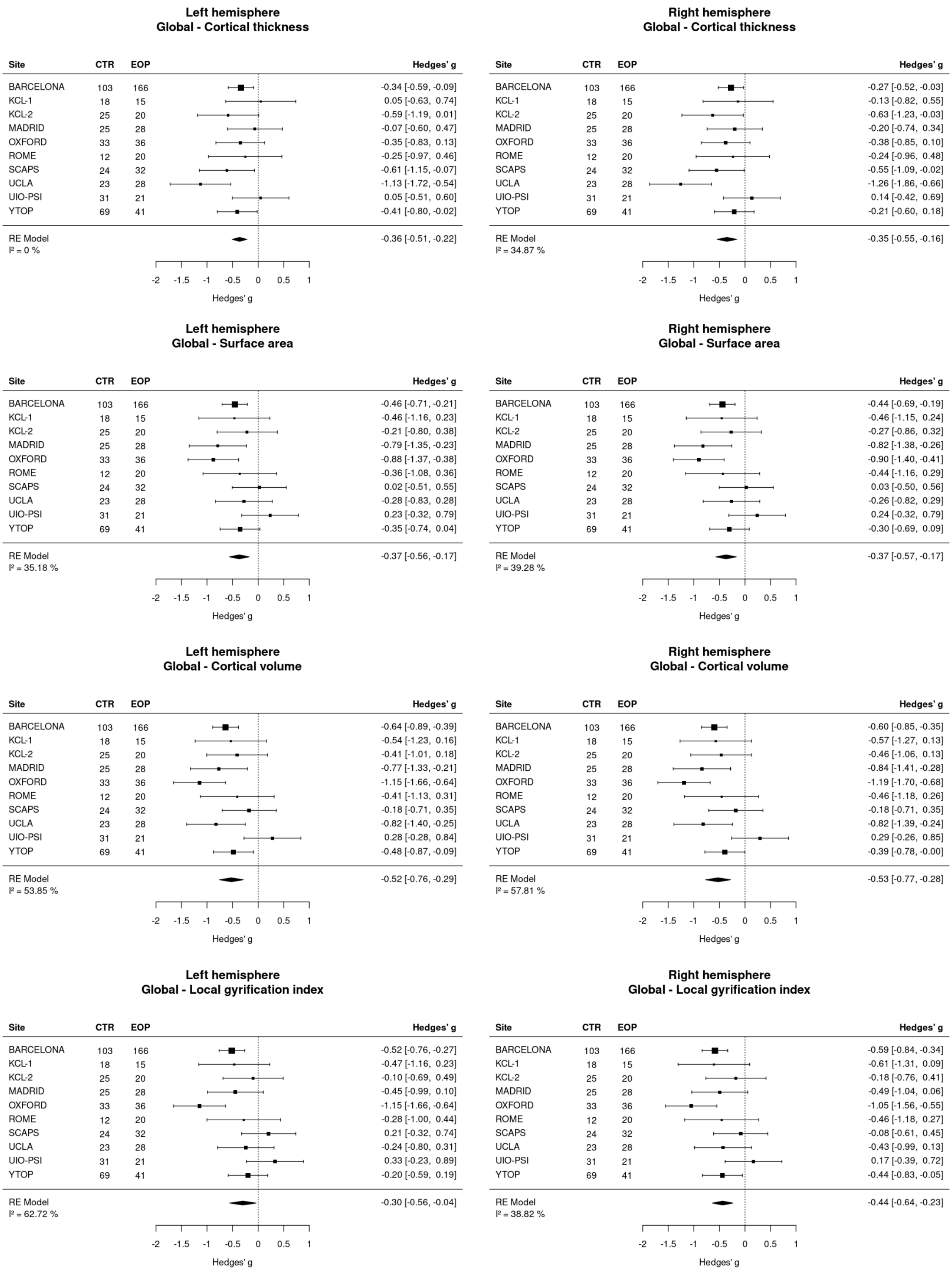


**Figure S7. Meta-analysis - Global cortical metrics.** Site-wise differences between early-onset psychosis (EOP) and healthy controls in global thickness, area, volume, and the Local Gyrification Index (LGI) for each hemisphere. Models were adjusted for age, age$^2$, and sex. Hedges' *g* effect sizes and 95% confidence intervals are shown for each site. The diamond indicates the pooled effect size and its confidence interval across sites.

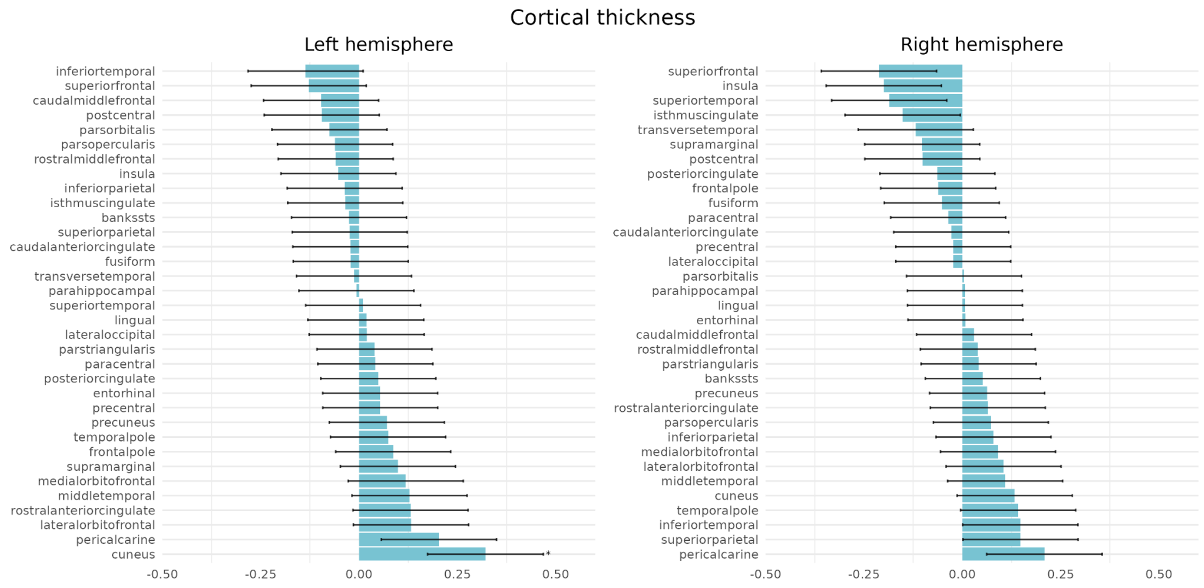


**Figure S8. Regional specificity - Cortical thickness.** Cohen's *d* effect sizes for comparisons between early-onset psychosis (EOP) and healthy controls adjusted for sex, age, and age$^2$ and global cortical thickness. Effect sizes are ranked by magnitude; significant differences after correction for multiple testing are marked with asterisks.

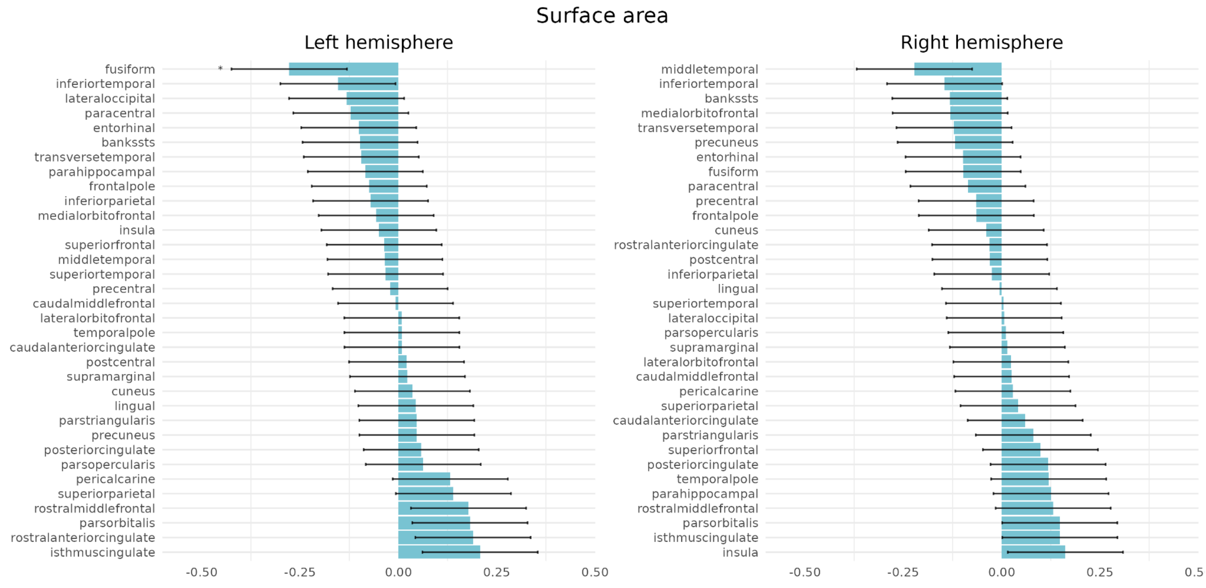


**Figure S9. Regional specificity - Surface area.** Cohen's *d* effect sizes for comparisons between early-onset psychosis (EOP) and healthy controls adjusted for sex, age, and age$^2$ and global surface area. Effect sizes are ranked by magnitude; significant differences after correction for multiple testing are marked with asterisks.

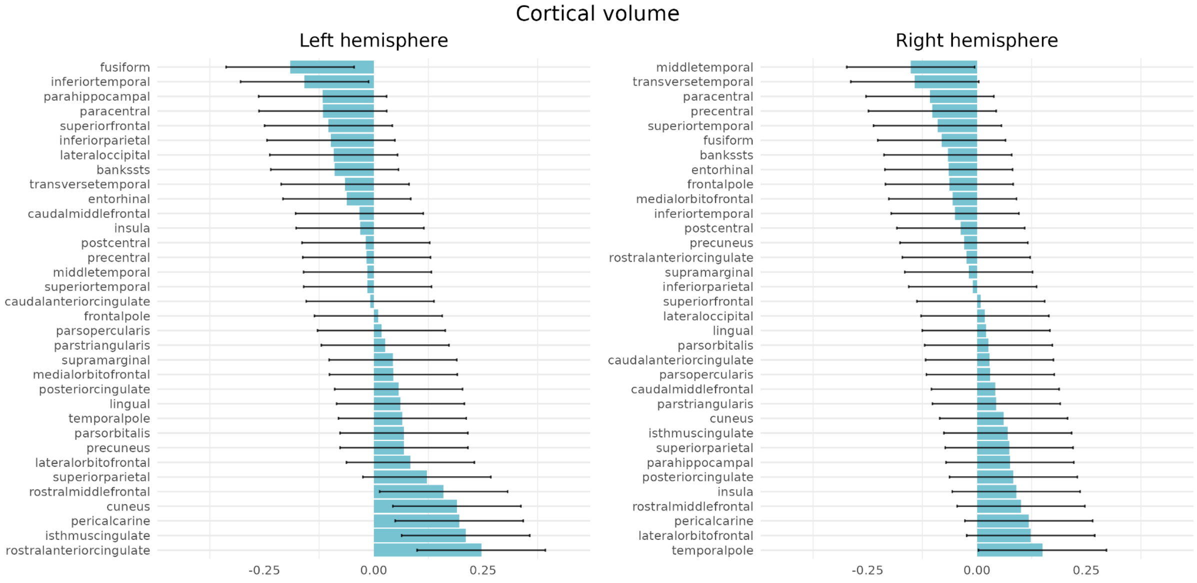


**Figure S10. Regional specificity - Cortical volume.** Cohen's *d* effect sizes for comparisons between early-onset psychosis (EOP) and healthy controls adjusted for sex, age, and age$^2$ and global cortical volume. Effect sizes are ranked by magnitude; none of the regions differed significantly after correction for multiple testing.

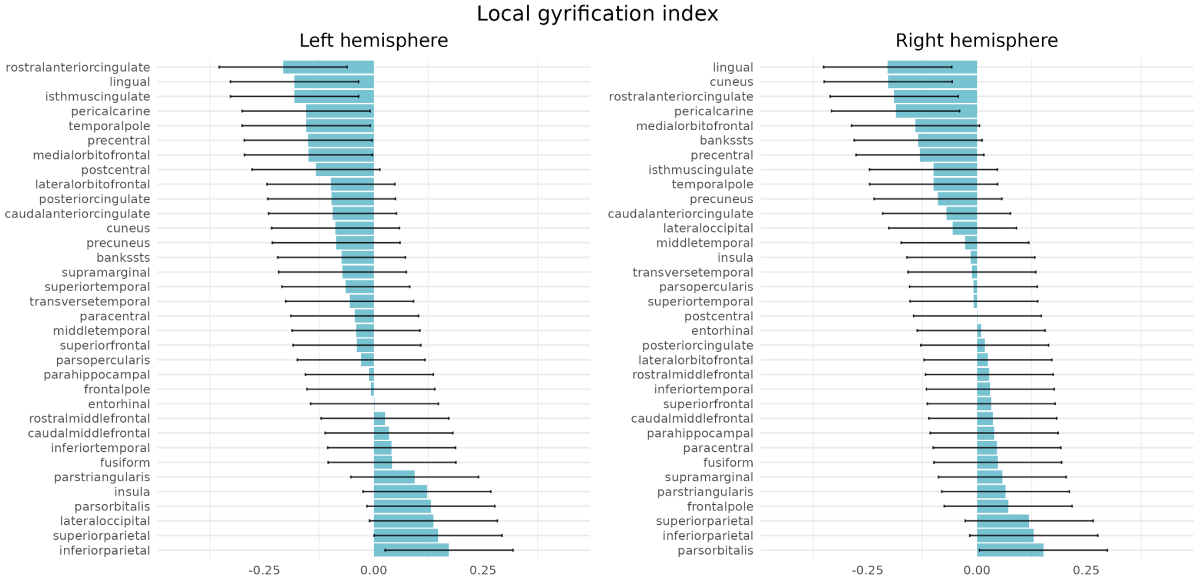


**Figure S11. Regional specificity - Local Gyrification Index (LGI).** Cohen's *d* effect sizes for comparisons between early-onset psychosis (EOP) and healthy controls adjusted for sex, age, and age$^2$ and global LGI. Effect sizes are ranked by magnitude; none of the regions differed significantly after correction for multiple testing.

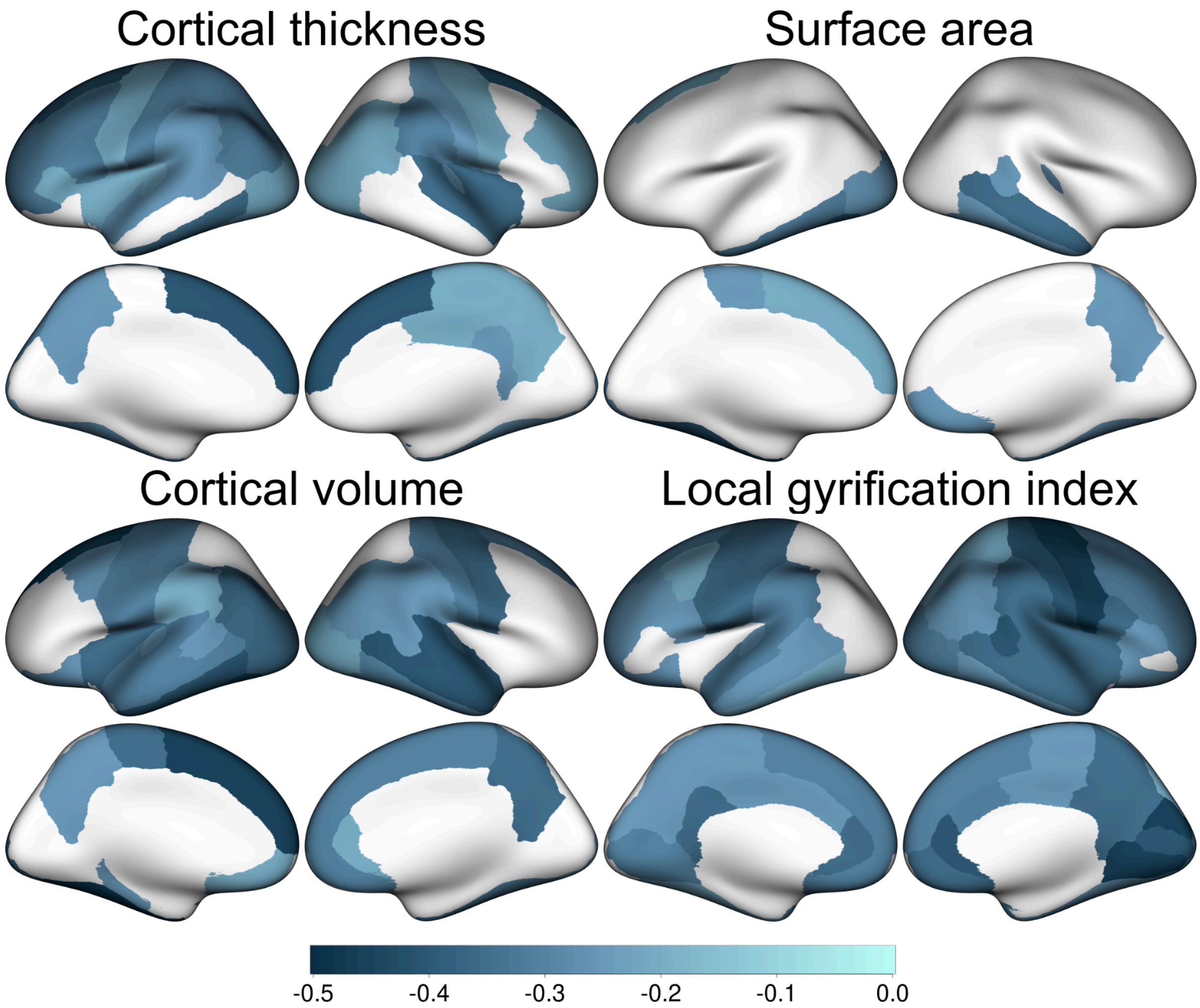


**Figure S12. Case-control differences adjusted for intracranial volume (ICV).** Cohen's *d* effect sizes for regions with significant differences in cortical thickness, surface area, cortical volume, and Local Gyrification Index (LGI) between early-onset psychosis (EOP) and healthy controls in models adjusted for sex, age, age$^2$, and ICV.

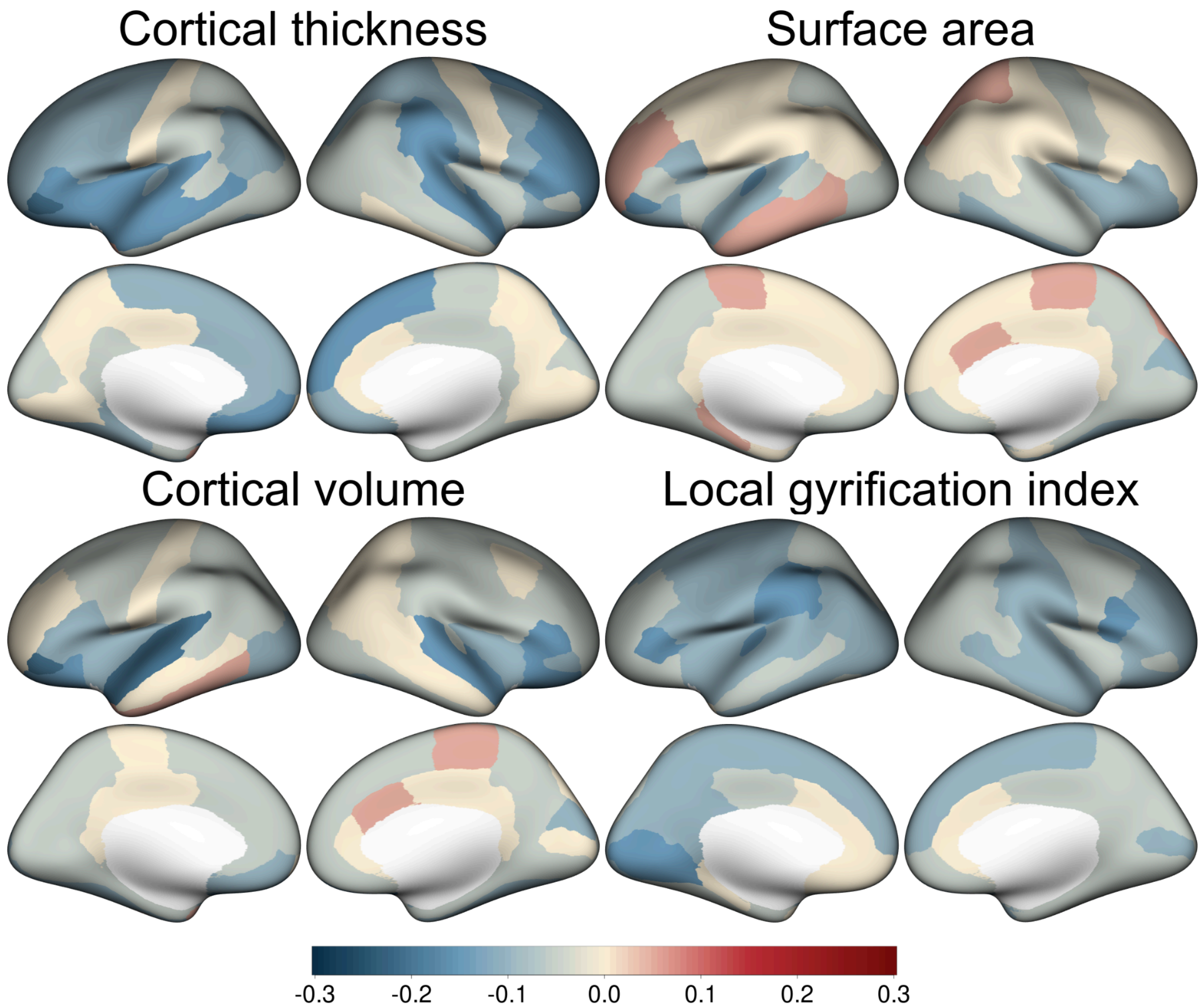


**Figure S13. Chlorpromazine-equivalent dose correlations.** Uncorrected partial correlations between chlorpromazine-equivalent antipsychotic medication dose and each cortical metric. Analyses were conducted for patients who were currently taking antipsychotic medication. No associatons survived correction for multiple testing.

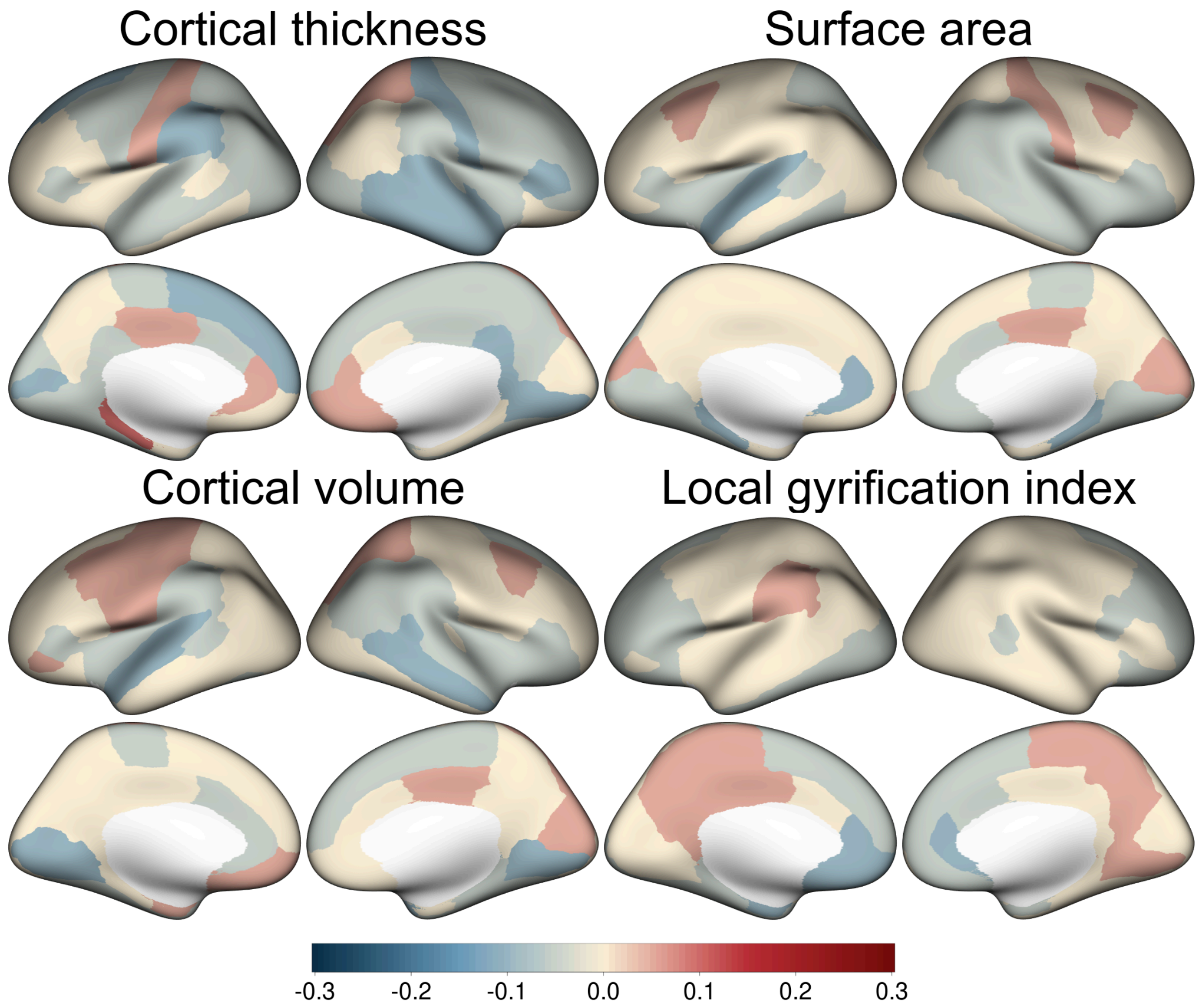


**Figure S14. PANSS positive symptom scores.** Uncorrected partial correlations between PANSS positive symptom scores and each cortical metric. No associatons survived correction for multiple testing.

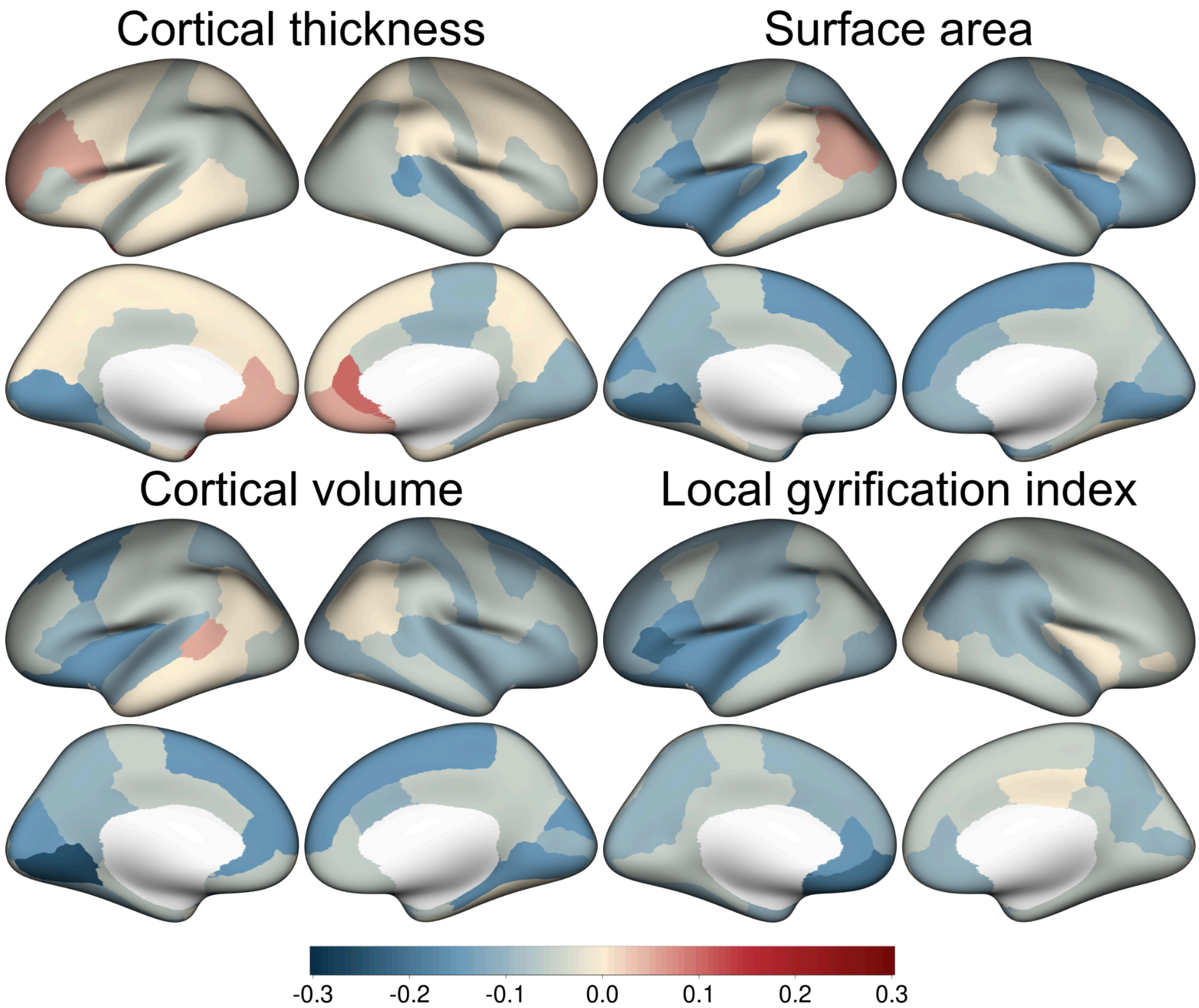


**Figure S15. PANSS negative symptom scores.** Uncorrected partial correlations between PANSS negative symptom scores and each cortical metric. Only the association with volume of the left lingual region was statistically significant (partial r=-0.21; $p_{FDR}$=0.014).

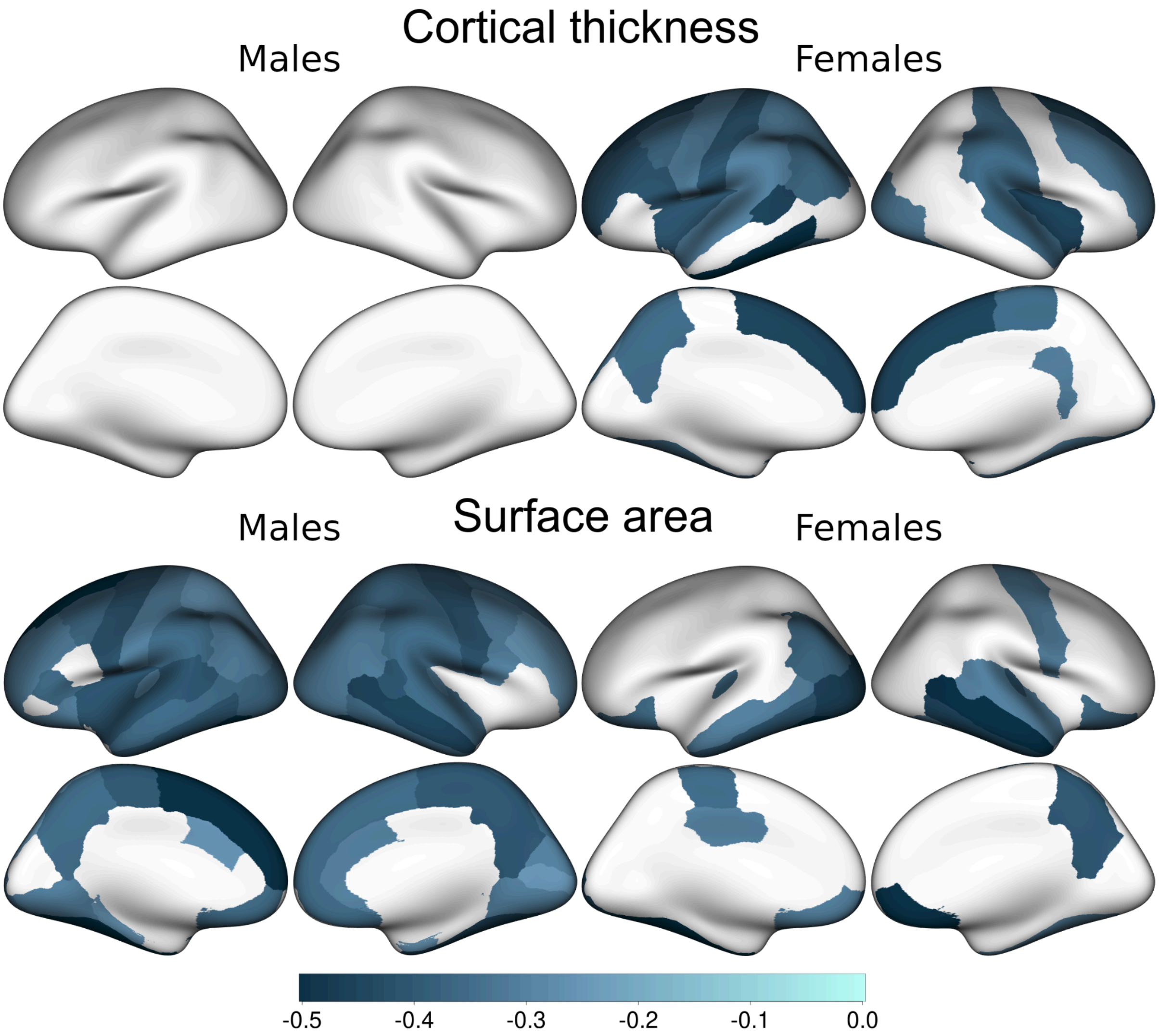


**Figure S16. Sex-stratified analyses - Cortical thickness and surface area.** Cohen's *d* effect sizes for regions with significant differences from comparisons between early-onset psychosis (EOP) and healthy controls using separate models for males and females.

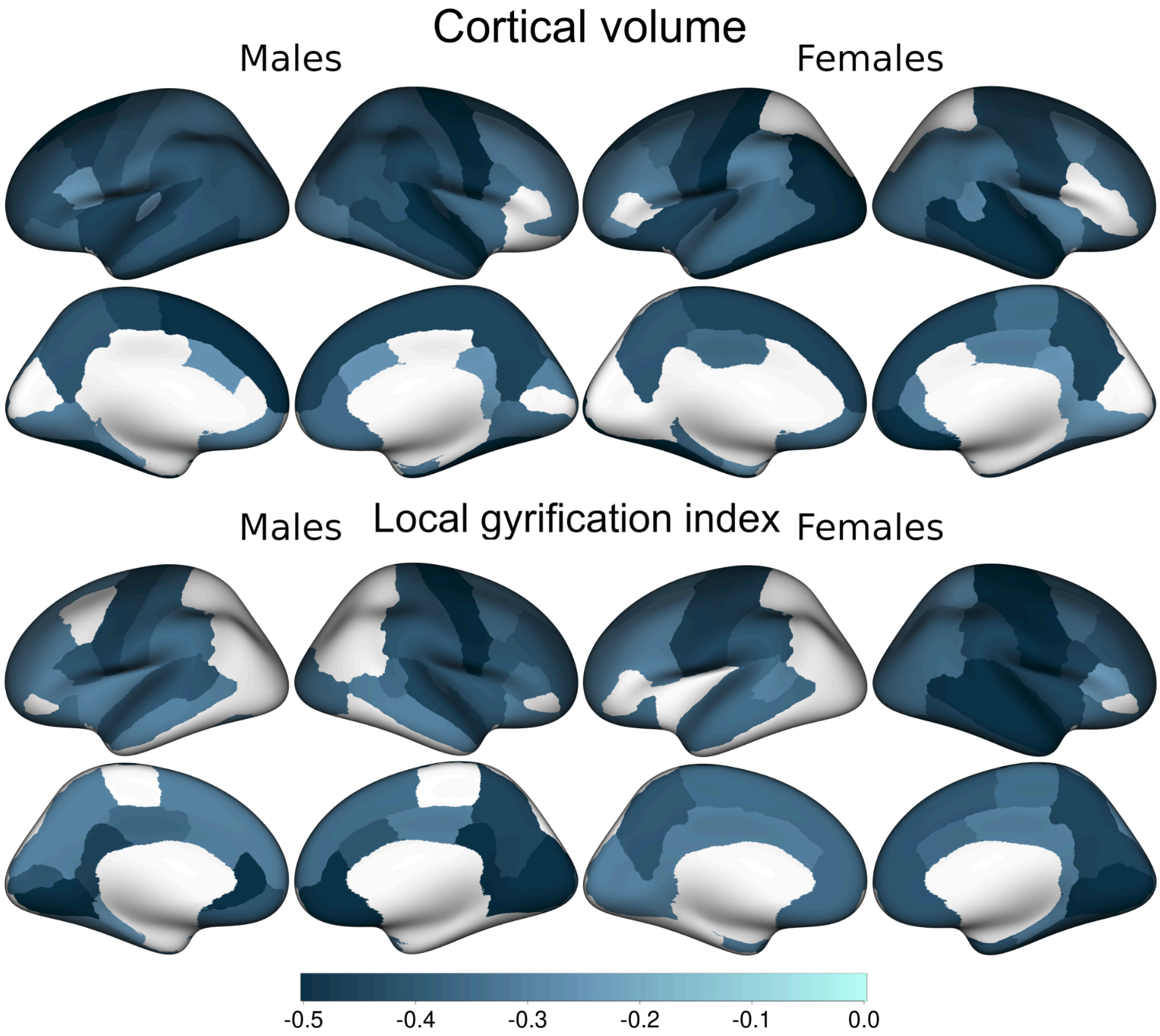


**Figure S17. Sex-stratified analyses - Cortical volume and Local Gyrification Index (LGI).** Cohen's *d* effect sizes for regions with significant differences from comparisons between early-onset psychosis (EOP) and healthy controls using separate models for males and females.

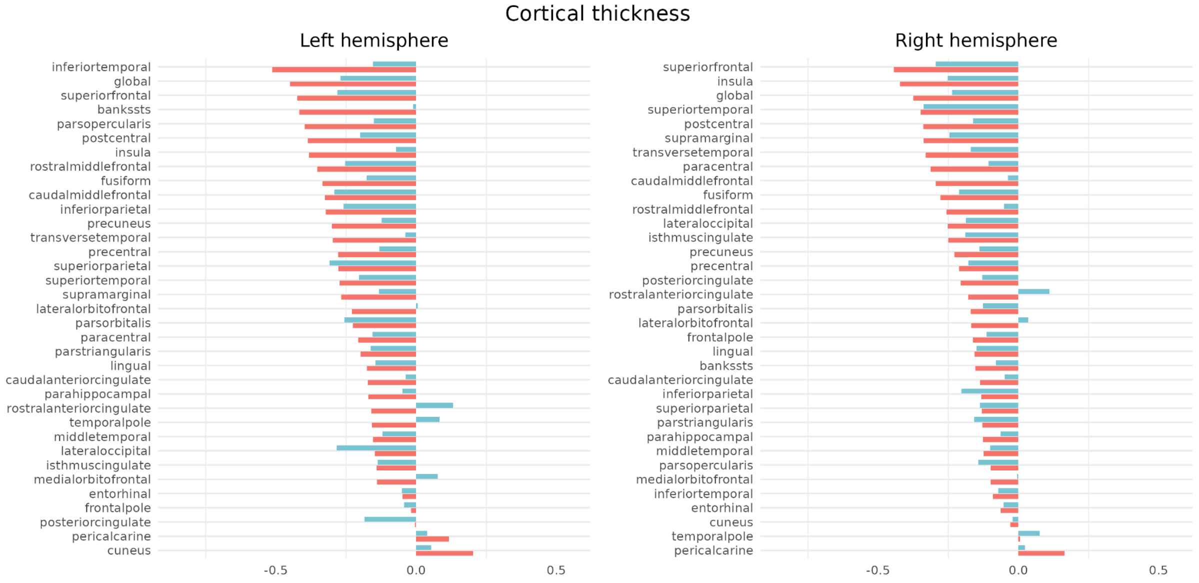


**Figure S18. Sex-stratified analyses - Cortical thickness.** Cohen's *d* effect sizes in the comparison between patients with early-onset psychosis (EOP) and healthy controls stratified by sex. Red bars indicate effect sizes for females and blue bars indicate effect sizes for males. Effect sizes are ordered by magnitude of effects for females.

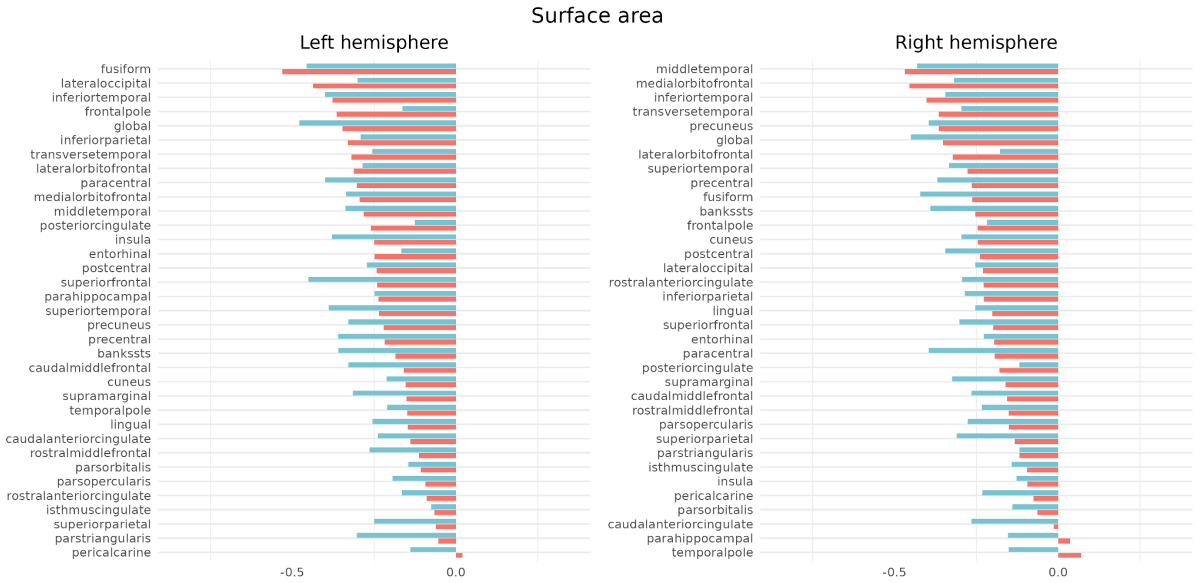


**Figure S19. Sex-stratified analyses - Surface area.** Cohen's *d* effect sizes in the comparison between patients with early-onset psychosis (EOP) and healthy controls stratified by sex. Red bars indicate effect sizes for females and blue bars indicate effect sizes for males. Effect sizes are ordered by magnitude of effects for females.

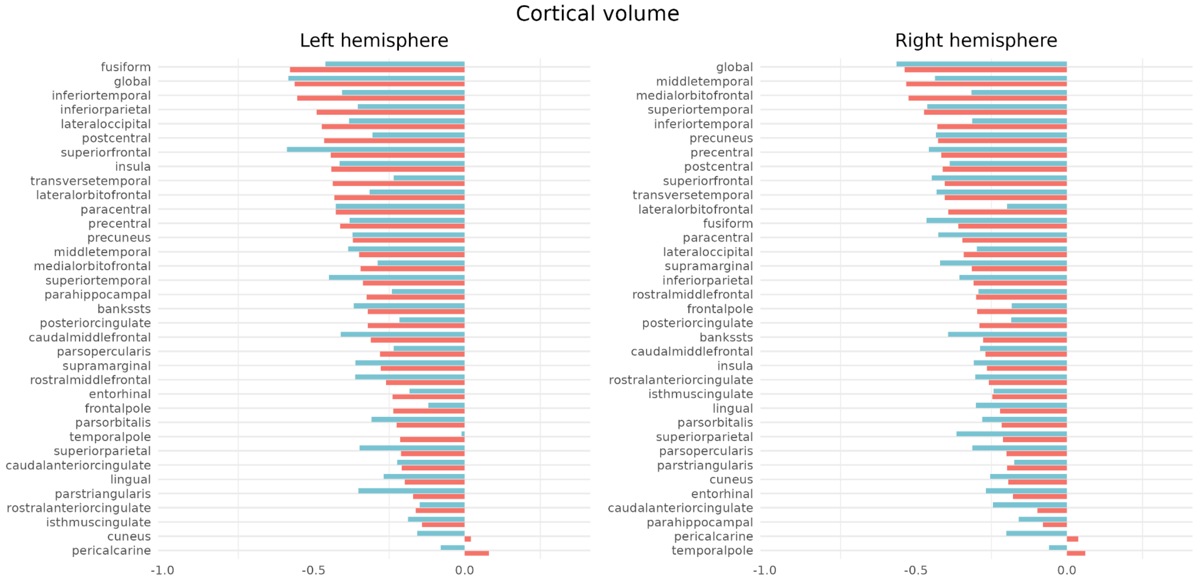


**Figure S20. Sex-stratified analyses - Cortical volume.** Cohen's *d* effect sizes in the comparison between patients with early-onset psychosis (EOP) and healthy controls stratified by sex. Red bars indicate effect sizes for females and blue bars indicate effect sizes for males. Effect sizes are ordered by magnitude of effects for females.

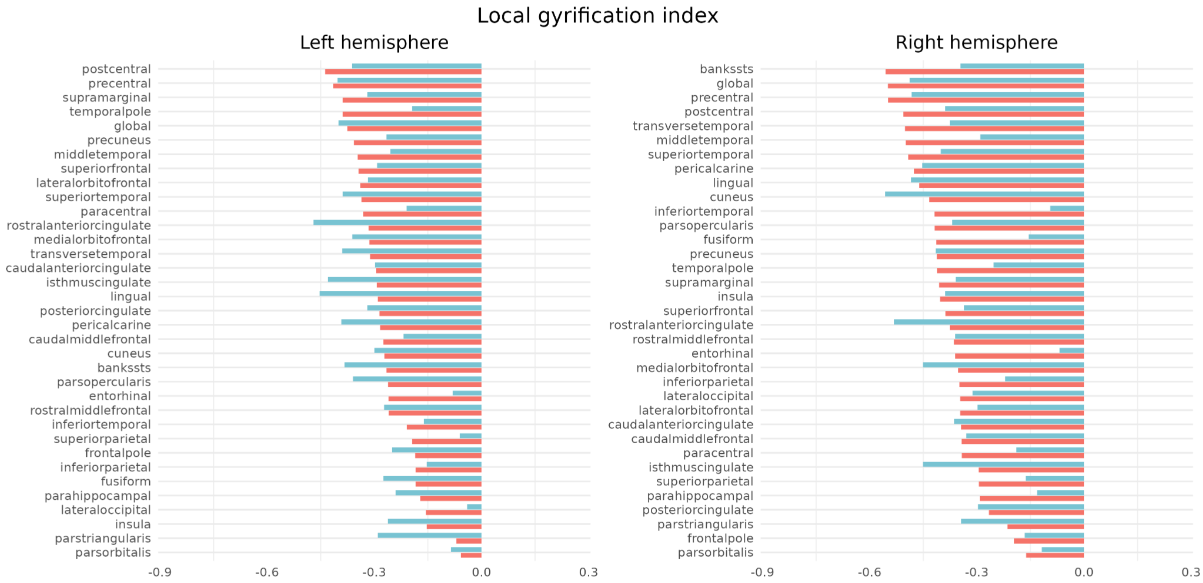


**Figure S21. Sex-stratified analyses - Local Gyrification Index (LGI).** Cohen's *d* effect sizes in the comparison between patients with early-onset psychosis (EOP) and healthy controls stratified by sex. Red bars indicate effect sizes for females and blue bars indicate effect sizes for males. Effect sizes are ordered by magnitude of effects for females.

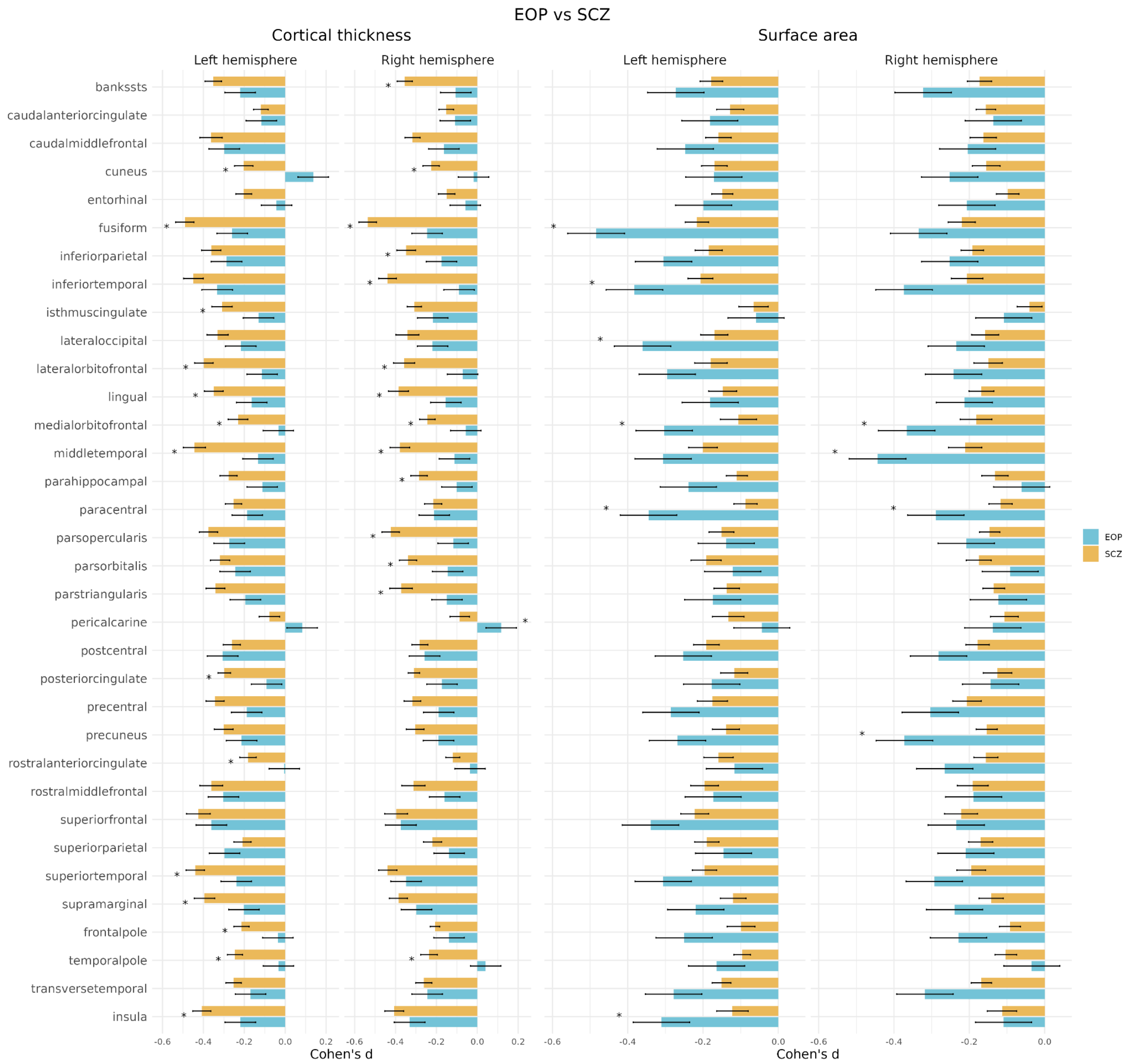


**Figure S22. Effect size comparison to adults with SCZ.** Cohen's *d* effect sizes from the case-control analyses for early-onset psychosis (EOP; *blue*) compared to those of the previous study on adults with schizophrenia (SCZ; *orange*). To harmonize covariates across studies, the regression models were only adjusted for age and sex. Significant differences in cortical thickness and surface area are marked with asterisks.

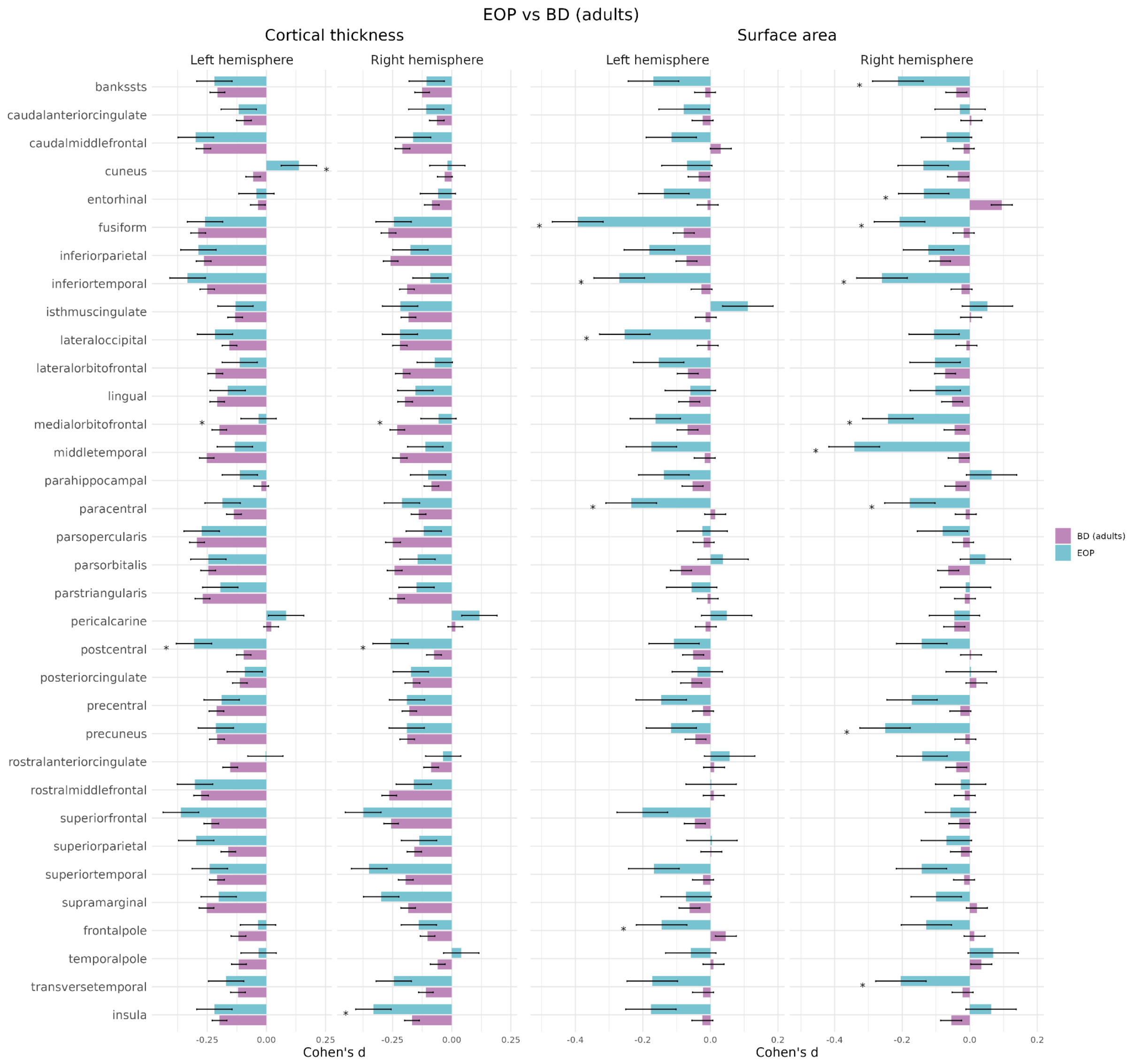


**Figure S23. Effect size comparison to adults with BD.** Cohen's *d* effect sizes from the case-control analyses for early-onset psychosis (EOP; *blue*) compared to those of the previous study on adults with bipolar disorders (BD; *purple*). To harmonize covariates across studies, the regression models for cortical thickness were only adjusted for age and sex, while surface area was also adjusted for intracranial volume (ICV). Significant differences in cortical thickness and surface area are marked with asterisks.

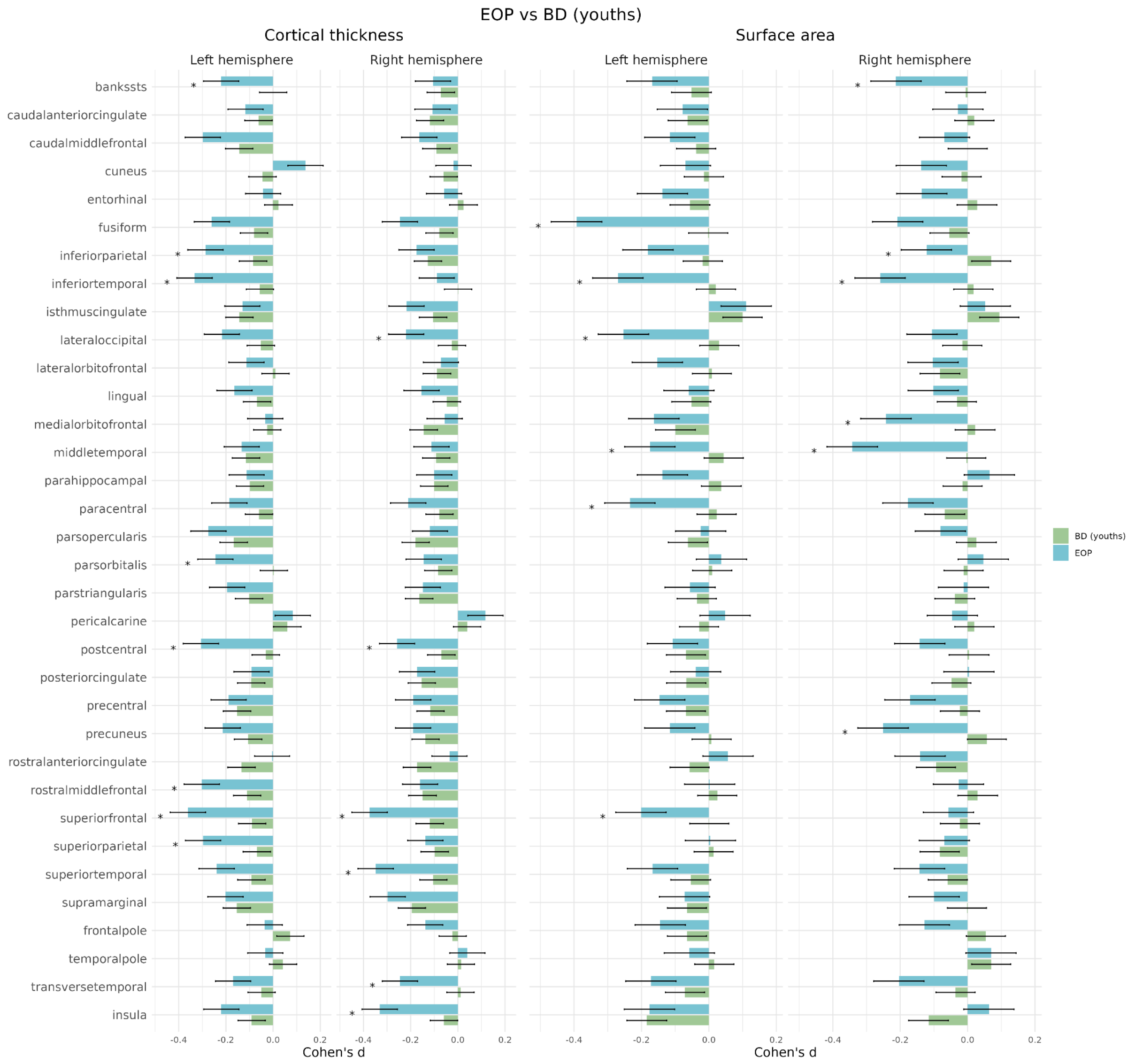


**Figure S24. Effect size comparison to youths with BD.** Cohen's *d* effect sizes from the case-control analyses for early-onset psychosis (EOP; *blue*) compared to those of the previous study on youths with bipolar disorders (BD; *green*). To harmonize covariates across studies, the regression models for cortical thickness were only adjusted for age and sex, while surface area was also adjusted for intracranial volume (ICV). Significant differences in cortical thickness and surface area are marked with asterisks.

### 3. Supplementary Tables

#### Table S1. Overview of participating research sites.

| Cohort | Principal investigator | Institution | City | Country | Data inclusion period |
|---|---|---|---|---|---|
| BARCELONA | Josefina Castro-Fornieles & Gisela Sugranyes | University of Barcelona | Barcelona | Spain | 2003 - 2020 |
| MADRID | Celso Arango | Hospital General Universitario Gregorio Marañón | Madrid | Spain | 2006 - 2014 |
| ROME | Fabrizio Piras | IRCCS Santa Lucia Foundation | Rome | Italy | 2012 - present |
| OXFORD | Anthony James | University of Oxford | Oxford | UK | 2005 - 2009 |
| KCL-1 | Sophia Frangou | King's College London | London | UK | 2002 - 2005 |
| KCL-2 | Anne-Kathrin J. Fett | King's College London | London | UK | 2010 - 2012 |
| UCLA | Carrie E. Bearden | University of California | Los Angeles | USA | 2011 - 2015 |
| SCAPS | Ingrid Agartz & Mathias Lundberg | Karolinska Institutet | Stockholm | Sweden | 2012 - 2019 |
| UIO-PSI | Bjørn Rishovd Rund & Lars T. Westlye | University of Oslo | Oslo | Norway | 2005 - 2007 |
| YTOP | Ingrid Agartz, Anne M. Myhre, and Nils Eiel Steen | University Hospital of Oslo | Oslo | Norway | 2012 - present |

#### Table S2. Recruitment procedures and criteria for each site.

| Cohort | Group | Clinical assessment | Recruitment information | Inclusion criteria | Exclusion criteria |
|---|---|---|---|---|---|
| BARCELONA | EOP | K-SADS; DSM-IV-TR | Referral from inpatient or outpatient units of the Department of Child and Adolescent Psychiatry and Psychology of the Hospital Clinic Barcelona | Age between 7-18 years; onset of first psychotic positive symptom within a psychotic episode before age of 18; diagnosis of a psychotic disorder per DSM-IV-TR criteria; written informed consent | Intellectual disability per DSM-IV-TR criteria (IQ < 70 & impaired functioning); pervasive developmental disorder; past history of head trauma with loss of consciousness; pregnancy |
| | CTR | K-SADS; DSM-IV-TR | Local catchment area via advertisements | Age between 7-18 years; written informed consent | Past history of psychotic illness; current diagnosis of any Axis-I DSM-IV-TR disorder; intellectual disability per DSM-IV-TR criteria (IQ < 70 & impaired functioning); past history of head trauma with loss of consciousness; pregnancy |
| MADRID | EOP | K-SADS; DSM-IV-TR | Referral from adolescent inpatient unit (Hospital General Universitario Gregorio Marañón) or local clinical services (PIENSA program) | Age between 7-18 years; onset of first psychotic positive symptom within a psychotic episode before age of 18; diagnosis of a psychotic disorder per DSM-IV-TR criteria; written informed consent | Intellectual disability per DSM-IV-TR criteria (IQ < 70 & impaired functioning); pervasive developmental disorder; past history of head trauma with loss of consciousness; pregnancy |
| | CTR | K-SADS; DSM-IV-TR | Local catchment area via advertisements | Age between 7-18 years; written informed consent | Past history of psychotic illness; current diagnosis of any Axis-I DSM-IV-TR disorder; intellectual disability per DSM-IV-TR criteria (IQ < 70 & impaired functioning); past history of head trauma with loss of consciousness; pregnancy |
| ROME | EOP | DSM-V psychiatric and personality disorders using the SCID-5-RV and SCID-5-PD | Local catchment area and referral from adolescent psychiatric units | Age between 10-18 years; onset of first psychotic positive symptom within a psychotic episode before age of 18; suitability for MRI scanning; written informed consent | A history of alcohol or drug abuse in the two years before the assessment; lifetime drug dependence; traumatic head injury with loss of consciousness; past or present major medical illness or neurological disorders; intellectual disability; pervasive developmental disorder |
| | CTR | DSM-V psychiatric and personality disorders using the SCID-5-RV and SCID-5-PD | Local catchment area via advertisements | Matched to patients; age between 10-18 years; suitability for MRI scanning; written informed consent | A history of alcohol or drug abuse in the two years before the assessment; lifetime drug dependence; traumatic head injury with loss of consciousness; past or present major medical illness or neurological disorders; any psychiatric disorder or intellectual disability |
| OXFORD | EOP | K-SADS-PL; DSM-IV | Local adolescent psychiatric units | Psychotic disorders included: schizophrenia | Moderate mental impairment; a history of substance abuse or pervasive developmental disorder; significant head injury; neurological disorder or major medical disorder |
| | CTR | KSADS-PL | Local general practitioners' practice | Healthy adolescents | Any medical/emotional/behavioral disorders; moderate mental impairment; a history of substance abuse or pervasive developmental disorder; significant head injury; neurological disorder or major medical disorder |
| KCL-1 | EOP | Structured Clinical Interview for DSM-IV (SCID) Axis I Disorders | Local clinical services | DSM IV Schizophrenia, 12-19 years old, onset of schizophrenia before age 18, no comorbid Axis I diagnosis, no mental retardation | History of head injury, current substance misuse, any medical condition, history of hereditary disease of the nervous system |
| | CTR | Structured Clinical Interview for DSM-IV (SCID) Axis I Disorders | Local area via advertisements | Age 12-19, no personal history of psychiatric disorders, no family history of psychosis in first-degree relatives | History of head injury, current substance misuse, any medical condition, history of hereditary disease of the nervous system |
| KCL-2 | EOP | ICD-10 diagnosed by treating clinician | Patients were recruited via consultant psychiatrists and via the Mental Health Research Network (MHRN) in South London and Maudsley, North East London, and South Essex Partnership University NHS Foundation Trusts | Age between 13-19 years; experienced a psychotic episode according to ICD-10 criteria, as diagnosed by their clinician; good command of the English language; being able and willing to give written informed consent | Diagnosed substance use/abuse or neurological conditions. |

| | | | | | |
|---|---|---|---|---|---|
| KCL-2 | CTR | No diagnosis confirmed in telephone screening | Control participants were recruited from local schools, the Institute of Psychiatry volunteer database 'Mindsearch', via colleagues and previous participants | Age between 13-19 years; good command of the English language; being able and willing to give written informed consent; no personal or family history of a psychotic illness | Diagnosed substance use/abuse or neurological conditions or a history of a psychiatric diagnosis and a family history of psychosis. |
| UCLA | EOP | DSM-IV. SCID Axis I Disorders | In-and outpatient clinics for child and adolescent mental health in the Greater Los Angeles area/website/local advertisements | Psychotic disorders included: schizophrenia spectrum disorder; age between 12-18 years; informed consent; no MRI contra-indications | Substance-induced psychotic disorder; IQ < 70; previous moderate to severe head injury; significant comorbid medial/neurological condition and/or history of head trauma with loss of consciousness |
| | CTR | DSM-IV. SCID Axis I Disorders | Local advertisements (online/brochures) in the Los Angeles areas | Matched to patients; age between 12-18 years; no history of major mental disorders; informed consent; no MRI contra-indications | History of mental health issues (contact with specialist services); first-degree relatives with a history of psychotic disorders; IQ < 70; significant comorbid medial/neurological condition and/or history of head trauma with loss of consciousness |
| SCAPS | EOP | DSM-IV | Specialist care unit of psychosis and bipolar disorder in the department of Child and Adolescent Psychiatry in Stockholm. Sweden | Psychotic disorders included: schizophrenia. schizoaffective disorder. psychotic depression; unspecified psychosis; bipolar I and II disorder; age between 12-18 years | Substance-induced psychotic disorder; IQ < 70; previous moderate to severe head injury; organic brain disease |
| | CTR | | Invitation by letter after random draw from the Swedish National Registry | Age between 12-18 years. good command of the Swedish language to complete interview and neurocognitive tests | History of mental health issues (contact with specialist services); previous or current use of psychotropic medication; first-degree relatives with a history of psychotic disorders; IQ < 70; previous moderate to severe head injury; organic brain disease |
| UIO-PSI | EOP | SCID-I, mod A-D; PANSS; GAF Split version | Recruited by clinicians at in- and outpatient child- and adolescent mental health clinics in Southern Norway | Age 12-18; broad schizophrenia spectrum disorder | Psychosis not otherwise specified (NOS); history of head injury; IQ < 70 |
| | CTR | Screened with MINI, screening mod | Recruited from schools in the patient catchment area, some from the database of the Norwegian Central Bureau of Statistics | | |
| YTOP | EOP | K-SADS-PL (2009)/ DSM-IV | In-and outpatient clinics for child and adolescent mental health in the greater Oslo area | Age between 12-18 years; diagnosis of psychotic disorder; good command of the Norwegian language to complete interview and neurocognitive tests | Substance-induced psychotic disorder; IQ < 70; previous moderate to severe head injury; organic brain disease |
| | CTR | K-SADS-PL (2009) | Invitation by letter after random draw from the Norwegian National Registry | Age between 12-18 years; good command of the Norwegian language to complete interview and neurocognitive tests | History of mental health issues (contact with specialist services); previous or current use of psychotropic medication; first-degree relatives with a history of psychotic disorders; IQ < 70; previous moderate to severe head injury; organic brain disease |

**Abbreviations:** EOP = early-onset psychosis; CTR = healthy controls; DSM = Structured Clinical Interview for Diagnostic and Statistical Manual of Mental Disorder; SCID = Structured Clinical Interview for DSM Disorders; K-SADS = Kiddie Schedule for Affective Disorders and Schizophrenia (PL = present and lifetime version); PANSS = Positive and Negative Syndrome Scale; IQ = intelligence quotient; MRI = magnetic resonance imaging.

### Table S3. Demographic and clinical information stratified by site.

| Cohort | Group | N | Age | Sex (Female) | Handedness (R/L/M) | IQ | FullDx (EOS/AFP/OTP) | PANSS Positive | PANSS Negative | AAO | DOI | CPZ | AP | LIT | AD | AE |
|---|---|---|---|---|---|---|---|---|---|---|---|---|---|---|---|---|
| BARCELONA | CTR | 103 | 16.3 [14.7, 17.4] | 58 (56.3%) | 25/6/0 | 104.0 [94.0, 110.8] | N.A. | N.A. | N.A. | N.A. | N.A. | N.A. | N.A. | N.A. | N.A. | N.A. |
| BARCELONA | EOP | 166 | 16.4 [15.1, 17.3] | 81 (48.8%) | 52/5/1 | 84.0 [76.0, 96.0] | 76/72/18 | 20 [16.0, 25.0] | 15 [10.5, 21.0] | 15.8 [14.6, 17.0] | 0.2 [0.1, 0.4] | 200.0 [150.0, 350.0] | 158 (95.2%) | 25 (15.1%) | 49 (29.5%) | 4 (2.4%) |
| MADRID | CTR | 25 | 16.0 [13.0, 17.0] | 12 (48.0%) | 20/2/0 | 110.0 [103.0, 119.0] | N.A. | N.A. | N.A. | N.A. | N.A. | N.A. | N.A. | N.A. | N.A. | N.A. |
| MADRID | EOP | 28 | 16.5 [14.0, 17.0] | 6 (21.4%) | 23/2/0 | 65.0 [54.0, 95.0] | 24/4/0 | 23.5 [19.8, 30.5] | 23 [16.0, 29.0] | 16.0 [14.8, 17.0] | 0.3 [0.2, 0.4] | NA | 28 (100.0%) | 0 | 2 (7.1%) | 1 (3.6%) |
| ROME | CTR | 12 | 13.0 [12.8, 14.2] | 7 (58.3%) | 10/2/0 | N.A. | N.A. | N.A. | N.A. | N.A. | N.A. | N.A. | N.A. | N.A. | N.A. | N.A. |
| ROME | EOP | 20 | 16.0 [15.0, 18.0] | 7 (35.0%) | 17/3/0 | N.A. | 16/0/4 | 11.5 [4.2, 20.8] | 27.5 [11.5, 42.5] | 15.0 [14.0, 17.0] | 1.0 [1.0, 1.2] | 200.0 [133.3, 283.3] | 18 (90.0%) | 0 | 3 (15.0%) | 3 (15.0%) |
| OXFORD | CTR | 33 | 16.3 [15.2, 17.2] | 18 (54.5%) | 29/4/0 | 109.0 [102.0, 121.0] | N.A. | N.A. | N.A. | N.A. | N.A. | N.A. | N.A. | N.A. | N.A. | N.A. |
| OXFORD | EOP | 36 | 16.6 [15.5, 17.0] | 17 (47.2%) | 28/6/1 | 85.5 [81.0, 100.2] | 36/0/0 | 23 [20.8, 24.2] | 16 [14.0, 18.0] | 14.7 [13.4, 15.7] | 1.4 [0.9, 2.4] | 300.0 [200.0, 412.0] | 36 (100.0%) | 0 | N.A. | 3 (8.3%) |
| KCL-1 | CTR | 18 | 16.0 [15.2, 17.2] | 7 (38.9%) | 16/2/0 | 100.5 [96.2, 104.5] | N.A. | N.A. | N.A. | N.A. | N.A. | N.A. | N.A. | N.A. | N.A. | N.A. |
| KCL-1 | EOP | 15 | 16.6 [15.9, 17.4] | 5 (33.3%) | 15/0/0 | 98.0 [93.5, 104.0] | 15/0/0 | 10 [7.5, 13.5] | 14 [12.0, 15.5] | 15.0 [14.1, 15.8] | 1.6 [1.0, 3.0] | N.A. | N.A. | N.A. | N.A. | N.A. |
| UCLA | CTR | 23 | 15.0 [13.0, 17.0] | 9 (39.1%) | 18/1/3 | 117.0 [107.0, 127.0] | N.A. | N.A. | N.A. | N.A. | N.A. | N.A. | N.A. | N.A. | N.A. | N.A. |
| UCLA | EOP | 28 | 16.0 [13.8, 17.0] | 15 (53.6%) | 25/1/2 | 105.0 [90.8, 113.5] | 17/7/4 | 22.5 [12.8, 32.8] | 26 [20.2, 39.8] | 13.0 [12.2, 14.8] | 1.0 [0.2, 2.8] | 200.0 [50.0, 325.0] | 16 (57.1%) | 1 (3.6%) | 14 (50.0%) | 2 (7.1%) |
| SCAPS | CTR | 24 | 16.9 [16.2, 17.7] | 19 (79.2%) | 22/2/0 | N.A. | N.A. | N.A. | N.A. | N.A. | N.A. | N.A. | N.A. | N.A. | N.A. | N.A. |
| SCAPS | EOP | 32 | 16.7 [15.6, 17.6] | 18 (56.2%) | 7/1/0 | N.A. | 2/19/11 | N.A. | N.A. | 15.2 [14.1, 16.6] | 1.0 [0.1, 2.4] | 166.5 [16.8, 258.5] | N.A. | N.A. | N.A. | N.A. |
| UIO-PSI | CTR | 31 | 15.0 [14.5, 17.0] | 16 (51.6%) | 28/3/0 | 108.0 [97.5, 116.5] | N.A. | N.A. | N.A. | N.A. | N.A. | N.A. | N.A. | N.A. | N.A. | N.A. |
| UIO-PSI | EOP | 21 | 16.0 [15.3, 17.0] | 12 (57.1%) | 19/2/0 | 98.0 [81.0, 109.0] | 14/0/7 | 13 [10.0, 16.5] | 13 [9.5, 15.0] | 15.0 [14.0, 16.0] | 1.2 [0.8, 2.2] | 67.0 [0.0, 300.0] | 10 (47.6%) | NA | 3 (14.3%) | 0 |
| YTOP | CTR | 69 | 16.2 [15.3, 17.4] | 38 (55.1%) | 65/4/0 | 107.0 [96.5, 114.0] | N.A. | N.A. | N.A. | N.A. | N.A. | N.A. | N.A. | N.A. | N.A. | N.A. |
| YTOP | EOP | 41 | 16.5 [15.7, 17.3] | 30 (73.2%) | 35/3/0 | 100.5 [88.0, 108.0] | 23/2/16 | 16 [14.0, 20.0] | 21 [14.0, 23.0] | 15.2 [13.0, 16.0] | 1.1 [0.7, 2.2] | 75.0 [0.0, 133.8] | 19 (46.3%) | 0 | 3 (7.3%) | 0 |

**Abbreviations:** EOP = early-onset psychosis; CTR = healthy controls; EOS = early-onset schizophrenia; AFP = affective psychosis; OTP = other psychotic disorders; PANSS = Positive and Negative Syndrome Scale; AAO = age at onset; DOI = duration of illness; AP = antipsychotic medication; CPZ = chlorpromazine-equivalent antipsychotic medication dose; LIT = lithium; AD = antidepressant medication; AE = antiepileptic medication; N.A. = not applicable. Handedness is given as number of right-handed, left-handed, and mixed individuals, among individuals where handedness information was available.

***Notes:*** The demographic and clinical information is given for the final mega-analysis dataset after 42 participants were excluded after quality control. The additional dataset used in the meta-analysis is described in **Table S4**.

#### Table S4. Demographic and clinical information for KCL-2 dataset.

| | CTR (N=25) | EOP (N=20) |
|---|---|---|
| **Age [years]** | 16.36 (1.6) | 16.95 (1.2) |
| **Sex [female]** | 0 (0%) | 0 (0%) |
| **Parental education [years]** | N.A. | N.A. |
| **Handedness [right]** | 25 (100.0%) | 20 (100.0%) |
| **IQ** | N.A. | N.A. |
| **Age at onset [years]** | N.A. | 16.3 (1.4) |
| **Duration of illness [years]** | N.A. | 1.1 (0.8) |
| **PANSS Positive** | N.A. | N.A. |
| **PANSS Negative** | N.A. | N.A. |
| **SAPS** | N.A. | N.A. |
| **SANS** | N.A. | N.A. |
| **AP use [yes]** | N.A. | 17 (85%) |
| **CPZ [mg/day]** | N.A. | N.A. |
| **Antiepileptic use [yes]** | N.A. | 3 (15%) |
| **Antidepressant use [yes]** | N.A. | 5 (25%) |
| **Lithium use [yes]** | N.A. | 0 (0%) |

**Abbreviations:** PANSS = Positive and Negative Syndrome Scale, AP = antipsychotic medication, CPZ = chlorpromazine-equivalent antipsychotic medication dose, N.A. = not applicable.

***Notes:*** Continuous variables are given as means (standard deviations) and categorical variables as counts (percentage of non-missing). This dataset was used only in the meta-analyses and information is reported for healthy controls (CTR) and the early-onset psychosis (EOP).

#### Table S5. MRI acquisition parameters for each site.

| Cohort | Scanner | Field strength | Scanner model | Sequence description | Acquisition direction | Resolution ($mm^3$) | TI (ms) | TE (ms) | TR (ms) | Flip angle (°) |
|---|---|---|---|---|---|---|---|---|---|---|
| BARCELONA | 1 | 1.5T | GE Genesis Signa | 3D T1-weighted | Axial | 1x1x1.5 | 300 | 5.168 | 12 | 20 |
| | 2 | 3T | Siemens TrioTim | MPRAGE sequence | Sagittal | 1x1x1 | 900 | 3.01 | 2300 | 9 |
| | 3 | - | - | - | - | - | - | - | - | - |
| MADRID | 4 | 1.5T | Philips | 3D T1-weighted | Sagittal | 1x0.94x0.94 | - | 9.2 | 25 | - |
| ROME | 5 | 3T | Philips Achieva | MPRAGE sequence | Sagittal | 0.5x0.5x0.9 | - | 5.3 | 11 | 8 |
| OXFORD | 6 | 1.5T | Siemens | 3D T1-weighted | Coronal | 1x1x1 | - | 5.6 | 12 | 19 |
| KCL-1 | 7 | 1.5T | GE | 3D T1-weighted | Coronal | 0.86x0.86x1.5 | 450 | 3.0 | 14 | 20 |
| KCL-2 | 8 | 3T | GE Signa | 3D T1-weighted | Coronal | 1.1x1.1x1.1 | - | 2.8 | 7 | 20 |
| UCLA | 9 | 3T | Siemens Tim Trio | 3D T1-weighted | Sagittal | 1x1x1 | 900 | 2.91 | 2300 | 9 |
| SCAPS | 10 | 3T | GE Discovery MR750 | BRAVO sequence | Sagittal | 1x1x1.2 | - | 3.06 | 7.9 | 12 |
| UIO-PSI | 11 | 1.5T | Siemens Sonata | 3D-SPGR sequence | Coronal | 1x1x1 | 1000 | 3.93 | 2730 | 7 |
| YTOP | 12 | 3T | GE Signa HDxt | FSPGR sequence | Sagittal | 1x1x1.2 | 450 | MinFull | 7.8 | 12 |
| | 13 | 3T | GE Discovery MR750 | BRAVO sequence | Sagittal | 1x1x1 | 450 | 3.18 | 8.16 | 12 |

**Abbreviations:** TI = inversion time; TE = echo time; TR = repetition time.

#### Table S6. Model summaries for global cortical metrics.

| | Left hemisphere | | | | | | | |
|---|---|---|---|---|---|---|---|---|
| | Global Cortical Thickness | | Global Surface Area | | Global Cortical Volume | | Global LGI | |
| ***Predictors*** | ***Estimates*** | ***p*** | ***Estimates*** | ***p*** | ***Estimates*** | ***p*** | ***Estimates*** | ***p*** |
| **Intercept** | 3.18 (-0.80 – 7.15) | 0.117 | -4.69 (-8.32 – -1.06) | 0.011 | -3.41 (-6.90 – 0.09) | 0.056 | 1.08 (-2.69 – 4.85) | 0.575 |
| **Age (years)** | -0.15 (-0.68 – 0.37) | 0.574 | 0.72 (0.24 – 1.20) | 0.003 | 0.67 (0.21 – 1.13) | 0.004 | 0.12 (-0.37 – 0.62) | 0.624 |
| **$Age^2$** | -0.00 (-0.02 – 0.02) | 0.81 | -0.02 (-0.04 – -0.01) | 0.004 | -0.03 (-0.04 – -0.01) | 0.001 | -0.01 (-0.03 – 0.01) | 0.245 |
| **Sex (female)** | -0.11 (-0.24 – 0.03) | 0.121 | -1.09 (-1.21 – -0.96) | <0.001 | -1.07 (-1.19 – -0.96) | <0.001 | -0.77 (-0.89 – -0.64) | <0.001 |
| **EOP** | -0.33 (-0.46 – -0.20) | <0.001 | -0.35 (-0.47 – -0.23) | <0.001 | -0.47 (-0.58 – -0.35) | <0.001 | -0.34 (-0.46 – -0.21) | <0.001 |
| **$R^2$ / $R^2$ adjusted** | 0.176 / 0.171 | | 0.314 / 0.310 | | 0.363 / 0.359 | | 0.259 / 0.255 | |
| | **Right hemisphere** | | | | | | | |
| | **Global Cortical Thickness** | | **Global Surface Area** | | **Global Cortical Volume** | | **Global LGI** | |
| ***Predictors*** | ***Estimates*** | ***p*** | ***Estimates*** | ***p*** | ***Estimates*** | ***p*** | ***Estimates*** | ***p*** |
| **Intercept** | 3.31 (-0.64 – 7.27) | 0.1 | -4.34 (-7.96 – -0.72) | 0.019 | -3.14 (-6.65 – 0.37) | 0.08 | -0.61 (-4.31 – 3.10) | 0.748 |
| **Age (years)** | -0.16 (-0.68 – 0.36) | 0.552 | 0.67 (0.19 – 1.14) | 0.006 | 0.63 (0.17 – 1.10) | 0.008 | 0.34 (-0.14 – 0.83) | 0.168 |
| **$Age^2$** | -0.00 (-0.02 – 0.01) | 0.791 | -0.02 (-0.04 – -0.01) | 0.007 | -0.02 (-0.04 – -0.01) | 0.002 | -0.02 (-0.03 – -0.00) | 0.044 |
| **Sex (female)** | -0.08 (-0.21 – 0.06) | 0.258 | -1.10 (-1.22 – -0.97) | <0.001 | -1.07 (-1.19 – -0.96) | <0.001 | -0.81 (-0.94 – -0.69) | <0.001 |
| **EOP** | -0.28 (-0.42 – -0.15) | <0.001 | -0.34 (-0.46 – -0.22) | <0.001 | -0.45 (-0.57 – -0.33) | <0.001 | -0.44 (-0.57 – -0.32) | <0.001 |
| **$R^2$ / $R^2$ adjusted** | 0.185 / 0.180 | | 0.317 / 0.313 | | 0.357 / 0.354 | | 0.285 / 0.281 | |

***Notes:*** Table summaries from the main model adjusted for sex, age, $age^2$ with diagnostic status (CTR/EOP) as the variable of interest. The outcome variables are mean-centered and scaled to facilitate comparison across cortical metrics.

**Table S7. Correlations between case-control alterations across diagnostic subgroups.**

**Cortical thickness**

| | EOS | AFP | OTP |
|---|---|---|---|
| **EOS** | 1 | 0.78 | 0.53 |
| **AFP** | 0.78 | 1 | 0.59 |
| **OTP** | 0.53 | 0.59 | 1 |

**Surface area**

| | EOS | AFP | OTP |
|---|---|---|---|
| **EOS** | 1 | 0.57 | 0.54 |
| **AFP** | 0.57 | 1 | 0.33 |
| **OTP** | 0.54 | 0.33 | 1 |

**Cortical volume**

| | EOS | AFP | OTP |
|---|---|---|---|
| **EOS** | 1 | 0.70 | 0.69 |
| **AFP** | 0.70 | 1 | 0.45 |
| **OTP** | 0.69 | 0.45 | 1 |

**Local Gyrification Index**

| | EOS | AFP | OTP |
|---|---|---|---|
| **EOS** | 1 | 0.58 | 0.60 |
| **AFP** | 0.58 | 1 | 0.68 |
| **OTP** | 0.60 | 0.68 | 1 |

***Notes:*** Pearson correlation coefficients for the Cohen's *d* effect sizes in the case-control analysis for the diagnostic subgroup analyses for cortical thickness, surface area, cortical volume, and the Local Gyrification Index (LGI).

#### Table S8. Statistical comparisons between correlation coefficients.

| Correlation 1 ($r_1$) | Correlation 2 ($r_2$) | $r_1$ | $r_2$ | z | p | |
|---|---|---|---|---|---|---|
| EOP vs SCZ - Cortical thickness | EOP vs BD (adults) - Cortical thickness | 0.62 | 0.61 | 0.24 | 0.809 | |
| EOP vs SCZ - Cortical thickness | EOP vs BD (youths) - Cortical thickness | 0.62 | 0.35 | 2.33 | 0.0199 | * |
| EOP vs BD (adults) - Cortical thickness | EOP vs BD (youths) - Cortical thickness | 0.61 | 0.35 | 2.4 | 0.0164 | * |
| EOP vs SCZ - Cortical thickness | EOP vs SCZ - Surface area | 0.62 | 0.49 | 1.16 | 0.247 | |
| EOP vs BD (adults) - Cortical thickness | EOP vs BD (adults) - Surface area | 0.61 | 0.12 | 3.4 | <0.001 | * |
| EOP vs BD (youths) - Cortical thickness | EOP vs BD (youths) - Surface area | 0.35 | 0.05 | 1.77 | 0.0763 | |
| EOP vs SCZ - Surface area | EOP vs BD (adults) - Surface area | 0.49 | 0.18 | 2.38 | 0.0172 | * |
| EOP vs SCZ - Surface area | EOP vs BD (youths) - Surface area | 0.49 | 0.05 | 2.76 | 0.00578 | * |
| EOP vs BD (adults) - Surface area | EOP vs BD (youths) - Surface area | 0.12 | 0.05 | 0.41 | 0.683 | |

***Notes:*** Statistical comparisons between Pearson correlation coefficients between early-onset psychosis (EOP) and adults with schizophrenia (SCZ), adults with bipolar disorders (BD), and youths with BD. The comparisons were conducted using Steiger's test and correlation coefficients that differed significantly are denoted with an asterisk.

#### Table S9. EOP vs SCZ - Cortical thickness. Significant effect size differences.

| | **Cortical thickness - EOP vs SCZ** | | | | | | |
|---|---|---|---|---|---|---|---|
| | **Left hemisphere** | | | | **Right hemisphere** | | |
| **Region** | **EOP** | **SCZ** | **p-value** | **Region** | **EOP** | **SCZ** | **p-value** |
| cuneus | 0.14 | -0.20 | $9.36 \times 10^{-5}$ | bankssts | -0.11 | -0.36 | $2.73 \times 10^{-3}$ |
| frontalpole | -0.04 | -0.21 | $3.21 \times 10^{-2}$ | cuneus | -0.02 | -0.23 | $1.42 \times 10^{-2}$ |
| fusiform | -0.26 | -0.49 | $7.98 \times 10^{-3}$ | fusiform | -0.25 | -0.54 | $7.60 \times 10^{-4}$ |
| insula | -0.22 | -0.41 | $2.98 \times 10^{-2}$ | inferiorparietal | -0.17 | -0.35 | $4.84 \times 10^{-2}$ |
| isthmuscingulate | -0.13 | -0.31 | $4.69 \times 10^{-2}$ | inferiortemporal | -0.09 | -0.44 | $4.90 \times 10^{-5}$ |
| lateralorbitofrontal | -0.11 | -0.40 | $1.07 \times 10^{-3}$ | lateralorbitofrontal | -0.07 | -0.36 | $1.63 \times 10^{-3}$ |
| lingual | -0.16 | -0.35 | $3.45 \times 10^{-2}$ | lingual | -0.15 | -0.39 | $9.33 \times 10^{-3}$ |
| medialorbitofrontal | -0.03 | -0.23 | $2.57 \times 10^{-2}$ | medialorbitofrontal | -0.06 | -0.24 | $2.47 \times 10^{-2}$ |
| middletemporal | -0.13 | -0.44 | $7.34 \times 10^{-4}$ | middletemporal | -0.11 | -0.38 | $2.59 \times 10^{-3}$ |
| posteriorcingulate | -0.09 | -0.30 | $1.06 \times 10^{-2}$ | parahippocampal | -0.10 | -0.29 | $2.87 \times 10^{-2}$ |
| rostralanteriorcingulate | 0.00 | -0.18 | $3.54 \times 10^{-2}$ | parsopercularis | -0.12 | -0.42 | $3.77 \times 10^{-4}$ |
| superiortemporal | -0.24 | -0.44 | $2.14 \times 10^{-2}$ | parsorbitalis | -0.14 | -0.34 | $2.33 \times 10^{-2}$ |
| supramarginal | -0.20 | -0.40 | $3.16 \times 10^{-2}$ | parstriangularis | -0.15 | -0.37 | $1.54 \times 10^{-2}$ |
| temporalpole | -0.03 | -0.25 | $1.04 \times 10^{-2}$ | pericalcarine | 0.12 | -0.09 | $2.22 \times 10^{-2}$ |
| | | | | temporalpole | 0.04 | -0.24 | $1.15 \times 10^{-3}$ |

***Notes:*** Regions where the Cohen's *d* effect sizes from the comparison of cortical thickness in early-onset psychosis (EOP) with healthy controls differed significantly from that of adults with schizophrenia (SCZ) from the previous large-scale study from ENIGMA-SZ.

#### Table S10. EOP vs SCZ - Surface area. Significant effect size differences.

| | Surface area - EOP vs SCZ | | | | | | |
|---|---|---|---|---|---|---|---|
| | Left hemisphere | | | | Right hemisphere | | |
| **Region** | **EOP** | **SCZ** | **p-value** | **Region** | **EOP** | **SCZ** | **p-value** |
| fusiform | -0.48 | -0.22 | $1.03{\times}10^{-3}$ | medialorbitofrontal | -0.37 | -0.18 | $3.21{\times}10^{-2}$ |
| inferiortemporal | -0.38 | -0.21 | $3.29{\times}10^{-2}$ | middletemporal | -0.44 | -0.21 | $7.67{\times}10^{-3}$ |
| insula | -0.31 | -0.12 | $2.83{\times}10^{-2}$ | paracentral | -0.29 | -0.12 | $3.39{\times}10^{-2}$ |
| lateraloccipital | -0.36 | -0.17 | $2.21{\times}10^{-2}$ | precuneus | -0.37 | -0.15 | $6.50{\times}10^{-3}$ |
| medialorbitofrontal | -0.30 | -0.11 | $2.67{\times}10^{-2}$ | | | | |
| paracentral | -0.34 | -0.09 | $1.49{\times}10^{-3}$ | | | | |

***Notes:*** Regions where the Cohen's *d* effect sizes from the comparison of surface area in early-onset psychosis (EOP) with healthy controls differed significantly from that of adults with schizophrenia (SCZ) from the previous large-scale study from ENIGMA-SZ.

#### Table S11. EOP vs adult BD - Cortical thickness. Significant effect size differences.

| | Cortical thickness - EOP vs BD (adults) | | | | | | |
|---|---|---|---|---|---|---|---|
| | Left hemisphere | | | | Right hemisphere | | |
| **Region** | **EOP** | **BD (adults)** | **p-value** | **Region** | **EOP** | **BD (adults)** | **p-value** |
| cuneus | 0.14 | -0.06 | $1.67{\times}10^{-2}$ | insula | -0.33 | -0.17 | $4.45{\times}10^{-2}$ |
| medialorbitofrontal | -0.03 | -0.20 | $4.00{\times}10^{-2}$ | medialorbitofrontal | -0.06 | -0.23 | $3.13{\times}10^{-2}$ |
| postcentral | -0.31 | -0.10 | $9.56{\times}10^{-3}$ | postcentral | -0.26 | -0.08 | $2.38{\times}10^{-2}$ |

***Notes:*** Regions where the Cohen's *d* effect sizes from the comparison of cortical thickness in early-onset psychosis (EOP) with healthy controls differed significantly from that of adults with bipolar disorders (BD) from the previous large-scale study from ENIGMA-BD.

#### Table S12. EOP vs adult BD - Surface area. Significant effect size differences.

| | Surface area - EOP vs BD (adults) | | | | | | |
|---|---|---|---|---|---|---|---|
| | Left hemisphere | | | | Right hemisphere | | |
| **Region** | **EOP** | **BD (adults)** | **p-value** | **Region** | **EOP** | **BD (adults)** | **p-value** |
| frontalpole | -0.14 | 0.05 | $1.84\times10^{-2}$ | bankssts | -0.21 | -0.04 | $3.27\times10^{-2}$ |
| fusiform | -0.39 | -0.08 | $1.14\times10^{-4}$ | entorhinal | -0.14 | 0.10 | $4.22\times10^{-3}$ |
| inferiortemporal | -0.27 | -0.03 | $2.61\times10^{-3}$ | fusiform | -0.21 | -0.02 | $1.89\times10^{-2}$ |
| lateraloccipital | -0.25 | -0.01 | $2.44\times10^{-3}$ | inferiortemporal | -0.26 | -0.02 | $3.61\times10^{-3}$ |
| paracentral | -0.23 | 0.01 | $2.15\times10^{-3}$ | medialorbitofrontal | -0.24 | -0.05 | $1.47\times10^{-2}$ |
| | | | | middletemporal | -0.34 | -0.03 | $1.40\times10^{-4}$ |
| | | | | paracentral | -0.18 | -0.01 | $4.10\times10^{-2}$ |
| | | | | precuneus | -0.25 | -0.01 | $3.31\times10^{-3}$ |
| | | | | transversetemporal | -0.20 | -0.02 | $2.37\times10^{-2}$ |

***Notes:*** Regions where the Cohen's *d* effect sizes from the comparison of surface area in early-onset psychosis (EOP) with healthy controls differed significantly from that of adults with bipolar disorders (BD) from the previous large-scale study from ENIGMA-BD.

#### Table S13. EOP vs BD (youths) - Cortical thickness. Significant effect size differences.

| | Cortical thickness - EOP vs BD (youths) | | | | | | |
|---|---|---|---|---|---|---|---|
| | Left hemisphere | | | | Right hemisphere | | |
| **Region** | **EOP** | **BD (youths)** | **p-value** | **Region** | **EOP** | **BD (youths)** | **p-value** |
| bankssts | -0.22 | 0.00 | $1.99\times10^{-2}$ | insula | -0.33 | -0.06 | $4.11\times10^{-3}$ |
| inferiorparietal | -0.29 | -0.09 | $3.28\times10^{-2}$ | lateraloccipital | -0.22 | -0.03 | $4.03\times10^{-2}$ |
| inferiortemporal | -0.33 | -0.06 | $3.49\times10^{-3}$ | postcentral | -0.26 | -0.07 | $4.71\times10^{-2}$ |
| parsorbitalis | -0.25 | 0.00 | $8.77\times10^{-3}$ | superiorfrontal | -0.37 | -0.12 | $7.20\times10^{-3}$ |
| postcentral | -0.31 | -0.03 | $3.69\times10^{-3}$ | superiortemporal | -0.35 | -0.10 | $9.96\times10^{-3}$ |
| rostralmiddlefrontal | -0.30 | -0.11 | $4.26\times10^{-2}$ | transversetemporal | -0.25 | 0.01 | $6.80\times10^{-3}$ |
| superiorfrontal | -0.36 | -0.09 | $4.12\times10^{-3}$ | | | | |
| superiorparietal | -0.30 | -0.07 | $1.62\times10^{-2}$ | | | | |

***Notes:*** Regions where the Cohen's *d* effect sizes from the comparison of cortical thickness in early-onset psychosis (EOP) with healthy controls differed significantly from that of youths (i.e., < 25 years) with bipolar disorders (BD) from the previous large-scale study from ENIGMA-BD.

#### Table S14. EOP vs BD (youths) - Surface area. Significant effect size differences.

| | **Surface area - EOP vs BD (youths)** | | | | | | |
|---|---|---|---|---|---|---|---|
| | **Left hemisphere** | | | | **Right hemisphere** | | |
| **Region** | **EOP** | **BD (youths)** | **p-value** | **Region** | **EOP** | **BD (youths)** | **p-value** |
| fusiform | -0.39 | 0.00 | $3.86 \times 10^{-5}$ | bankssts | -0.21 | -0.01 | $2.99 \times 10^{-2}$ |
| inferiortemporal | -0.27 | 0.02 | $2.13 \times 10^{-3}$ | inferiorparietal | -0.12 | 0.07 | $4.20 \times 10^{-2}$ |
| lateraloccipital | -0.25 | 0.03 | $2.66 \times 10^{-3}$ | inferiortemporal | -0.26 | 0.02 | $3.47 \times 10^{-3}$ |
| middletemporal | -0.17 | 0.04 | $2.06 \times 10^{-2}$ | medialorbitofrontal | -0.24 | 0.02 | $5.50 \times 10^{-3}$ |
| paracentral | -0.23 | 0.02 | $6.52 \times 10^{-3}$ | middletemporal | -0.34 | 0.00 | $3.62 \times 10^{-4}$ |
| superiorfrontal | -0.20 | 0.00 | $3.22 \times 10^{-2}$ | precuneus | -0.25 | 0.06 | $1.14 \times 10^{-3}$ |

***Notes:*** Regions where the Cohen's *d* effect sizes from the comparison of surface area in early-onset psychosis (EOP) with healthy controls differed significantly from that of youths (i.e., < 25 years) with bipolar disorders (BD) from the previous large-scale study from ENIGMA-BD.
